# Population-based multi-horizon machine-learning prognostication for prostate adenocarcinoma: 10-year competing-risk and race-stratified validation

**DOI:** 10.64898/2026.11.24.26368666

**Authors:** Junho Song, Jaime Herrera-Caceres, Jay D. Raman, Julian Chavarriaga Soto

## Abstract

**Background:** Prostate cancer risk tools estimate cancer-specific mortality but do not account for the competing risk of death from other causes. It remains unknown whether machine learning improves the joint prediction of prostate cancer-specific and other-cause mortality. It is also unclear whether excluding race and ethnicity improves model fairness.

**Methods:** We included men diagnosed with prostate adenocarcinoma in 2010–2015 using complete clinical records from the US Surveillance, Epidemiology, and End Results program. Discrete-time competing-risk models (four machine-learning algorithms and spline regression) jointly predicted 1–10-year prostate cancer-specific (PCSM), other-cause (OCM) and all-cause mortality from clinical stage, grade group, PSA, biopsy-core involvement, age and social variables. Using cross-validation and a temporal split, we assessed discrimination, calibration and net benefit against five established tools and compared subgroup performance with and without race and ethnicity.

**Findings:** Among 130 634 men (median follow-up 11 years [IQR 9–12]), 9575 died of prostate cancer and 24 952 of other causes. At 10 years, XGBoost AUCs were 0·872 (PCSM), 0·742 (OCM) and 0·795 (all-cause). Gains over the best cancer risk tool were 0·034, 0·128 and 0·087, respectively. Regression performed similarly (AUC difference ≤0·005). Calibration was good overall and by race and ethnicity. Excluding race and ethnicity left discrimination unchanged but underpredicted OCM in Black men (observed-to-expected ratio 1·16 [1·13–1·19]) and overpredicted it in Asian or Pacific Islander men (0·80 [0·76–0·85]). At the 10-year life-expectancy threshold, 2·1% and 3·8% of these men, respectively, were reclassified. Excluding competing deaths from training overestimated 10-year PCSM risk by 36%.

**Interpretation:** Modeling competing risks mattered more than algorithm choice. In this retrospective analysis, excluding race and ethnicity worsened calibration for other-cause mortality in Black, Asian or Pacific Islander and Hispanic men. Prognostic tools should report both outcomes and assess calibration by race and ethnicity.

**Funding:** This study received no funding.

**Research in context:** *Evidence before this study:* We searched PubMed on Sept 25, 2026, without language or date restrictions, for studies of machine learning or artificial intelligence for mortality or survival prediction in prostate cancer that referred to competing, other-cause, non-cancer or cancer-specific outcomes (46 records). We screened titles and read the abstracts of potentially relevant records. Previous registry-based studies predicted a single end point, usually 10-year prostate cancer-specific mortality or overall and cancer-specific survival modeled separately. Most reported concordance indices, which do not directly assess predicted risks at a fixed time, and compared algorithms with categorical risk groups rather than with a flexible regression model. One study compared random forests with multinomial regression for cause of death at a single horizon.^1^ We found no study that jointly modeled prostate cancer and other-cause mortality across several horizons with evaluation that accounted for censoring and competing risks, or that reported calibration by race and ethnicity together with the effect of excluding race as a predictor.

*Added value of this study:* In a large population-based cohort, a single discrete-time competing-risk model estimated 1-, 3-, 5- and 10-year risks of prostate cancer death, other-cause death and all-cause death. It was well calibrated overall, over time and across racial and ethnic groups. Four machine-learning algorithms and a spline-based regression model performed similarly (AUC differences <0·01). All five models discriminated better than the clinical tools for prostate cancer mortality. The larger gains for other-cause and all-cause mortality were expected because the tools were designed to stratify cancer risk. Excluding race and ethnicity from the model left discrimination essentially unchanged but led to underprediction of other-cause mortality in Black men and overprediction in Asian or Pacific Islander and Hispanic men. Among men with non-metastatic intermediate or high-risk disease, up to one in 20 crossed the threshold used in this study to approximate 10-year life expectancy. Among reclassified men, observed mortality favored the model that included race and ethnicity only in the Asian or Pacific Islander subgroup. Ignoring competing deaths when labeling outcomes, as is common in machine-learning studies, overestimated prostate cancer mortality risk. Adjustment for tumor characteristics at diagnosis largely accounted for higher prostate cancer mortality in Black men. Their higher other-cause mortality persisted after adjustment.

*Implications of all the available evidence:* Prognostic counseling at diagnosis should present prostate cancer and competing mortality together, over horizons that match treatment decisions. In this study, accounting for competing deaths had more effect on predicted risk than changing the algorithm. Studies of prognostic tools should report calibration by race and ethnicity and assess how it changes when those variables are excluded. Before clinical use, external validation with stage, biopsy core and comorbidity data is needed.

## Introduction

Prostate cancer is the most frequently diagnosed cancer in men in the USA and a leading cause of cancer death worldwide, with the number of cases projected to double by 2040.^2–4^ Most men are diagnosed with non-metastatic disease, and in contemporary cohorts, 15-year prostate cancer mortality is low even with initial monitoring.^5^ For most men, death from other causes is far more likely.^6^ Guidelines therefore recommend weighing the risk of cancer progression against life expectancy when choosing between surveillance and treatment.^7–9^ The tools most widely used at diagnosis (the D’Amico classification, the UCSF Cancer of the Prostate Risk Assessment [CAPRA] score, NCCN risk groups, the Cambridge Prognostic Groups [CPG] and ISUP grade groups) were developed to stratify cancer risk. None estimates the risk of dying from other causes.^10–15^ Individualized multivariable models that estimate both outcomes, such as PREDICT Prostate, have improved discrimination but depend on inputs that are not always available.^16,17^

Machine learning has been used to predict outcomes in prostate cancer.^18,19^ Yet systematic reviews show no consistent benefit over logistic regression for clinical prediction.^20^ Three features of the current literature limit interpretation. First, most studies model a single outcome at a single horizon. Second, studies often censor or exclude men who die of other causes. This can overestimate the absolute risk of prostate cancer death.^21,22^ Third, discrimination is typically summarized with a concordance index, which does not directly assess predicted risk at a fixed time, and calibration is rarely reported.^23,24^ Performance across sociodemographic groups is also seldom examined, although algorithms can encode or amplify inequities.^25–28^ This gap is particularly important in prostate cancer, given the higher risks of both prostate cancer-specific and other-cause mortality among Black men.^29,30^

Following TRIPOD+AI,^31^ we developed and internally validated models that use information available at diagnosis to jointly predict prostate cancer-specific, other-cause and all-cause mortality at 1, 3, 5 and 10 years. We compared four machine-learning algorithms with a flexible regression model and with five established risk-stratification tools. We evaluated discrimination, calibration and clinical utility overall, over time and across racial and ethnic groups, including the effect of excluding race and ethnicity from the models. We also quantified racial and ethnic differences in cause-specific mortality. Finally, we implemented the regression model as a web calculator.

## Methods

### Study design and data source

We conducted a population-based retrospective cohort study using the SEER program of the US National Cancer Institute.^32,33^ SEER collects demographic, tumor, first-course treatment and vital-status data from population-based cancer registries, with cause of death derived from death certificates and classified by the SEER cause-specific death algorithm.^34^ Follow-up extended through Dec 31, 2023. Reporting follows TRIPOD+AI^31^ (checklist in the appendix).

### Ethics and consent

This secondary analysis used de-identified SEER Research Data accessed under the National Cancer Institute data-use agreement. The study involved no contact with participants or access to identifiable private information. The National Cancer Institute does not require institutional review board approval for access to standard SEER Research Data. Individual informed consent was not obtained.

### Participants

We included men aged 20 years or older with microscopically confirmed acinar adenocarcinoma of the prostate (International Classification of Diseases for Oncology, 3rd edition, code 8140) as their first primary malignancy, diagnosed from 2010 to 2015. Death-certificate-only and autopsy-only cases were excluded. County attributes, treatment, sequence number and single-year age came from a second export of the same SEER database, linked record by record (appendix). We selected this period because clinical T, N and M categories and biopsy-core counts were coded uniformly under Collaborative Staging. Standardized PSA and Gleason pattern fields were available from 2010, and the period allowed at least 8 years of potential follow-up. For the primary analysis, we excluded deaths of unknown cause and men without a biopsy Gleason score, pretreatment PSA, clinical T category, regional node or distant metastasis status, or biopsy-core counts (figure 1). Characteristics of excluded men are shown in the appendix.

**Figure 1:**
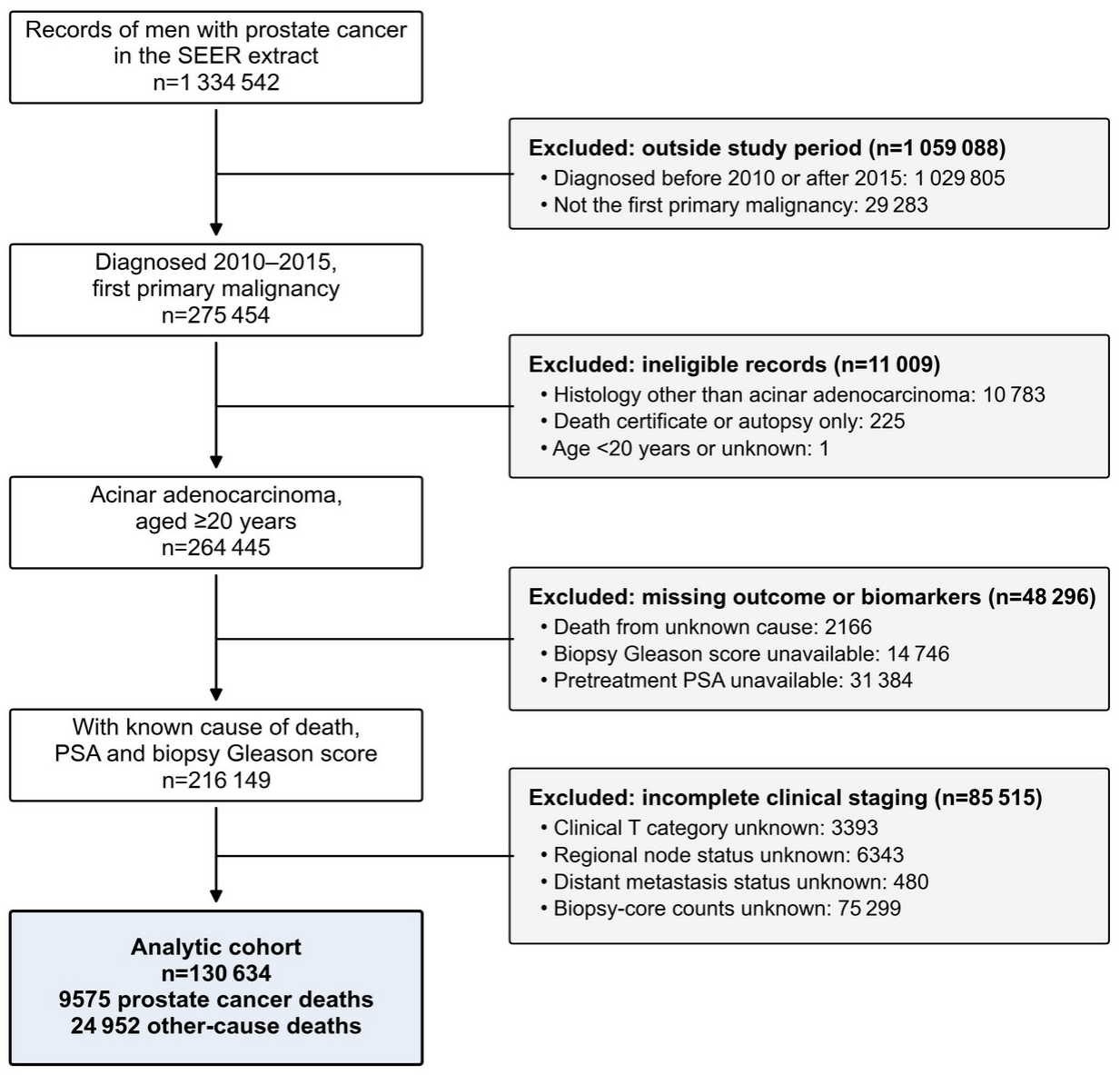
Study flow diagram. SEER=Surveillance, Epidemiology, and End Results. PSA=prostate-specific antigen.

### Predictors

Candidate predictors were prespecified as variables that are routinely known at diagnosis, are recorded consistently in population registries, and are established prognostic factors^12,16,17^: age, serum PSA (ng/mL, with values recorded as ≥98 ng/mL set to 98), biopsy grade group (from the clinical Gleason patterns), clinical T category, regional node (cN) status, distant metastasis (M0, M1a, M1b or M1c), the percentage of positive biopsy cores, race and ethnicity, marital status, county-level median household income (inflation-adjusted to 2024) and rurality (Rural–Urban Continuum Code), both time-dependent county attributes for the year of diagnosis. Race and ethnicity (non-Hispanic White, non-Hispanic Black, Hispanic, non-Hispanic Asian or Pacific Islander, non-Hispanic American Indian or Alaska Native, unknown) were recorded by registries from medical records. We treated these categories as social constructs that may reflect exposure to racism and differences in access to care.^26,27^ Pathological Gleason score and treatment were not used in the primary model because they are unknown when the prognosis is first discussed. First-course radical prostatectomy, radiotherapy and chemotherapy were added in a secondary, treatment-augmented model. To quantify the value of the clinical inputs, a secondary registry-minimal model used only age, PSA, biopsy Gleason score and the social variables, in the same men. We excluded date of diagnosis because trends over time might not apply to future patients.

### Outcomes

The outcomes were prostate cancer-specific mortality (PCSM), other-cause mortality (OCM) and all-cause mortality at 1, 3, 5 and 10 years after diagnosis. Death attributed to prostate cancer by the SEER cause-specific death classification defined PCSM. Deaths attributed to other causes defined OCM. Prostate cancer death and death from other causes are competing events because each precludes the other. The estimands were therefore the cumulative incidence functions (absolute risks) of each cause, which sum to the risk of all-cause death.^21^ We used survival time in months from SEER. We counted deaths within 12h months of diagnosis as events by horizon h and censored men who were alive at their last contact.

### Model development

We used a discrete-time competing-risk formulation.^35^ Each man contributed one record per year of follow-up (up to 10 years), labeled as survival, death from prostate cancer, or death from other causes during that year. Censored intervals contributed information until last contact. A multinomial classifier estimated the cause-specific hazard of each event in each interval given the predictors and the interval index. Absolute risks were obtained as CIFₖ(h)=Σⱼ≤h S(j−1)λₖ(j), where λₖ(j) is the predicted hazard of cause k in interval j and S(j)=Πₘ≤ⱼ[1−λ₁(m)−λ₂(m)]. A single fitted model yields absolute risks that cannot decrease over time. At each horizon, all-cause mortality risk equals the sum of the two cause-specific risks.

We compared five models: XGBoost,^36^ LightGBM,^37^ histogram-based gradient boosting, random forest, and multinomial logistic regression. We used flexible multinomial logistic regression as the reference model.^20^ It included restricted cubic splines (four knots) for age and log-PSA, restricted cubic splines (three knots) for the percentage of positive cores, indicators for grade group, clinical T category (T1, T2a, T2 not otherwise specified, T2b–T2c, T3a, T3b–T4), cN1, M1 with M1b and M1c contrasts, and categorical social variables, interval-specific intercepts (a flexible baseline hazard), and interactions of follow-up time with age, log-PSA, grade group and M1 to allow time-varying effects. Boosting hyperparameters (tree depth or number of leaves, and minimum node size) were chosen from a small prespecified grid by multinomial log-loss on an inner 10% split of the development data, and the number of trees was set by early stopping. Selected values were: XGBoost, maximum depth 4, minimum child weight 100; LightGBM, maximum leaves 15, minimum leaf size 50; and histogram gradient boosting, maximum leaves 15, minimum leaf size 500 (learning rate 0·05 throughout). The random forest used 300 trees, a minimum leaf size of 100 and a 25% bootstrap sample per tree. We did not use class reweighting or resampling to change outcome frequencies. XGBoost was prespecified as the primary machine-learning model based on preliminary analyses and because gradient boosting captures non-linear effects and interactions and can handle missing values directly. We report performance for all five models. We implemented the regression model, which has an explicit formula, as a web calculator that reproduces its predictions exactly (see Data sharing).

### Comparator risk-stratification tools

We compared the models with UCSF-CAPRA,^11,12^ NCCN risk groups,^7^ CPG,^13^ the D’Amico classification^10^ and ISUP grade group,^14^ each computed with its published inputs: PSA, grade group from the clinical Gleason patterns, clinical T category and, for CAPRA, age and the percentage of positive cores (definitions in the appendix). We combined NCCN groups into low, intermediate and high (including very high risk, cN1 and M1). We assessed discrimination using the ordered categories of each tool. For calibration and decision-curve analysis, each category was assigned its observed (censoring-weighted) risk in the training folds, so that each tool was recalibrated on the training data.

### Validation and performance measures

Internal validation used five-fold cross-validation with patient-level folds stratified by outcome and diagnosis period. For cross-validation, we calculated performance measures from out-of-fold predictions and tuned the models within the training folds. For the temporal split within SEER, we trained models on men diagnosed in 2010–2012 and evaluated 1-, 3- and 5-year predictions in men diagnosed in 2013–2015.

We used inverse probability of censoring weights estimated by the Kaplan–Meier method to account for censoring before each prediction horizon.^38^ Discrimination was measured by the time-dependent AUC under the competing-risk definition, in which cases are men who died of the cause of interest by the horizon and controls are all others, including men who died of the competing cause.^23,38^ Overall accuracy was measured by the Brier score and the index of prediction accuracy (IPA, calculated as 1 − model Brier score / null-model Brier score).^39^ Calibration was assessed with the observed-to-expected (O:E) ratio (observed and mean predicted risk both computed in the censoring-weighted population with evaluable status at the horizon), calibration intercept (calibration-in-the-large) and slope, and flexible calibration curves (weighted logistic regression of the outcome on restricted cubic splines of the logit prediction). These curves were summarized by the integrated calibration index (ICI) and E90.^40^ Clinical utility was assessed by decision-curve analysis, with net benefit computed from censoring-weighted true and false positives.^41,42^ We obtained 95% CIs, and paired differences between models, from 500 patient-level bootstrap resamples (200 for subgroup, temporal and sensitivity analyses), re-estimating the censoring distribution in each resample. We checked the performance measures using simulated competing-risk data with known outcomes (appendix).

### Fairness analysis

Following recommendations for algorithmic auditing,^27,28^ we estimated discrimination (AUC), O:E ratio, calibration slope and ICI separately in each racial and ethnic group at every horizon, using the cross-validated predictions. To assess the effect of excluding race and ethnicity,^26^ we refitted the XGBoost and regression models without race and ethnicity and compared subgroup calibration and discrimination with the models including race and ethnicity. We prespecified three analyses to assess the implications of differences in calibration. First, because guidelines use a life expectancy of at least 10 years to select men for curative treatment,^7,9^ we used a predicted 10-year all-cause mortality of 50% or higher as a proxy for life expectancy below 10 years. Within each racial and ethnic group, we cross-tabulated classifications from the two models. A predicted 10-year all-cause mortality of 50% corresponds to a predicted median survival of 10 years. Guideline-based life expectancy estimates use health-adjusted life tables and require comorbidity data, which SEER lacks.^7,9^ Second, for men whose classification changed, we compared each model’s mean predicted risk with censoring-weighted observed 10-year mortality. Third, we examined the consistency of the calibration shift after excluding race and ethnicity across strata of age, NCCN group, income, rurality, marital status and diagnosis period, and estimated the difference in net benefit per 1000 men at the life-expectancy threshold.

### Disparities analysis

To examine racial and ethnic differences in mortality, we estimated cumulative incidence of PCSM and OCM by race and ethnicity with the Aalen–Johansen estimator. We then fitted Cox proportional hazards models for cause-specific hazards,^21^ with sequential adjustment. Model 1 adjusted for age. Model 2 added tumor characteristics (PSA, biopsy grade group, clinical T category, cN, M and the percentage of positive cores). Model 3 added marital status, income and rurality, and model 4 added first-course treatment.

### Sensitivity analyses

We examined whether the discrete-time model was as accurate as horizon-specific (’direct’) XGBoost classifiers. These were trained with other-cause deaths classified as non-events and with inverse probability of censoring weights to account for censoring. We also quantified the bias of excluding men who died of other causes before the prediction horizon from training. We assessed the incremental value of treatment variables in the treatment-augmented model. Finally, because missing biopsy-core counts were the largest exclusion, we repeated the main analyses in all men with known clinical T, N and M category, using the clinical model without the percentage of positive cores, and compared the two models within the men of the primary cohort.

### Sample size and missing data

The sample size was fixed by the available data. With 9575 PCSM and 24 952 OCM events, the cohort exceeded the estimated minimum sample size for a model with fewer than 60 parameters.^43^ The analysis was complete case for the clinical variables (PSA, biopsy Gleason score and grade group, clinical T, N and M category and biopsy-core counts). Missing social variables were modeled as an explicit unknown category (regression) or handled natively by the tree algorithms.

### Software

Analyses used Python 3.13 with scikit-learn 1.8, XGBoost 3.2, LightGBM 4.6, lifelines 0.30 and SHAP 0.51 (TreeSHAP for model explanation^44^). The complete analysis code, which regenerates every number, table and figure in this report from the SEER extract, is available from the authors (see Data sharing).

## Results

Of 1 334 542 records, 130 634 men met the eligibility criteria (figure 1). Median age was 65 years (IQR 59–70) and median PSA was 6·7 ng/mL (IQR 4·9–10·5). The cohort comprised 88 226 White (67·5%), 20 968 Black (16·1%), 12 674 Hispanic (9·7%), 6977 Asian or Pacific Islander (5·3%) and 568 American Indian or Alaska Native (0·4%) men (table 1). During a median follow-up of 11 years (IQR 9–12), 34 527 men died, including 9575 of prostate cancer and 24 952 of other causes. Prostate cancer therefore accounted for 27·7% of deaths. The 10-year cumulative incidence was 6·9% (6·8– 7·1) for PCSM and 16·9% (16·7–17·1) for OCM. Other-cause death was the more common outcome in 11 of 16 strata of age and biopsy grade group (1, 2–3, 4 and 5). Prostate cancer death predominated only in men younger than 55 years with grade group 4 and in men with grade group 5 at any age (appendix).

**Table 1:** Characteristics of the cohort, overall and by race and ethnicity.

| Characteristic | Overall | Non-Hispanic White | Non-Hispanic Black | Hispanic | Non-Hispanic Asian or Pacific Islander | Non-Hispanic American Indian or Alaska Native |
|---|---|---|---|---|---|---|
| Patients, n | 130 634 | 88 226 | 20 968 | 12 674 | 6977 | 568 |
| Age, years, median (IQR) | 65 (59–70) | 65 (60–71) | 62 (57–68) | 65 (58–70) | 66 (61–72) | 65 (60–70) |
| Age <55 | 14 519 (11·1%) | 8473 (9·6%) | 3625 (17·3%) | 1672 (13·2%) | 535 (7·7%) | 62 (10·9%) |
| Age 55–64 | 47 812 (36·6%) | 31 472 (35·7%) | 8844 (42·2%) | 4599 (36·3%) | 2232 (32·0%) | 198 (34·9%) |
| Age 65–74 | 51 641 (39·5%) | 36 343 (41·2%) | 6855 (32·7%) | 4766 (37·6%) | 2971 (42·6%) | 238 (41·9%) |
| Age ≥75 | 16 662 (12·8%) | 11 938 (13·5%) | 1644 (7·8%) | 1637 (12·9%) | 1239 (17·8%) | 70 (12·3%) |
| PSA, ng/mL, median (IQR) | 6·7 (4·9–10·5) | 6·4 (4·8–9·9) | 7·2 (5·1–12·7) | 7·1 (5·1–11·8) | 7·6 (5·4–12·5) | 8·0 (5·4–14·6) |
| PSA <10 | 94 887 (72·6%) | 66 418 (75·3%) | 13 893 (66·3%) | 8704 (68·7%) | 4574 (65·6%) | 349 (61·4%) |
| PSA 10–20 | 21 299 (16·3%) | 13 478 (15·3%) | 3816 (18·2%) | 2274 (17·9%) | 1436 (20·6%) | 120 (21·1%) |
| PSA >20 | 14 448 (11·1%) | 8330 (9·4%) | 3259 (15·5%) | 1696 (13·4%) | 967 (13·9%) | 99 (17·4%) |
| **Biopsy grade group** |  |  |  |  |  |  |
| 1 | 53 611 (41·0%) | 36 685 (41·6%) | 7937 (37·9%) | 5526 (43·6%) | 2596 (37·2%) | 218 (38·4%) |
| 2 | 34 933 (26·7%) | 23 639 (26·8%) | 6134 (29·3%) | 3132 (24·7%) | 1604 (23·0%) | 142 (25·0%) |
| 3 | 16 847 (12·9%) | 11 235 (12·7%) | 2901 (13·8%) | 1527 (12·0%) | 978 (14·0%) | 68 (12·0%) |
| 4 | 13 655 (10·5%) | 8865 (10·0%) | 2259 (10·8%) | 1367 (10·8%) | 1016 (14·6%) | 47 (8·3%) |
| 5 | 11 588 (8·9%) | 7802 (8·8%) | 1737 (8·3%) | 1122 (8·9%) | 783 (11·2%) | 93 (16·4%) |
| Positive biopsy cores, %, median (IQR) | 33 (17–58) | 33 (17–57) | 36 (17–64) | 33 (17–60) | 33 (17–54) | 38 (17–67) |
| **Clinical T stage** |  |  |  |  |  |  |
| T1 | 89 688 (68·7%) | 58 890 (66·7%) | 15 992 (76·3%) | 8794 (69·4%) | 4846 (69·5%) | 363 (63·9%) |
| T2a | 8496 (6·5%) | 6273 (7·1%) | 934 (4·5%) | 689 (5·4%) | 477 (6·8%) | 43 (7·6%) |
| T2b–T2c | 9156 (7·0%) | 6496 (7·4%) | 1135 (5·4%) | 910 (7·2%) | 504 (7·2%) | 33 (5·8%) |
| T2, NOS | 18 389 (14·1%) | 13 199 (15·0%) | 2250 (10·7%) | 1763 (13·9%) | 841 (12·1%) | 92 (16·2%) |
| T3a | 4037 (3·1%) | 2843 (3·2%) | 488 (2·3%) | 417 (3·3%) | 246 (3·5%) | 29 (5·1%) |
| T3b–T4 | 868 (0·7%) | 525 (0·6%) | 169 (0·8%) | 101 (0·8%) | 63 (0·9%) | 8 (1·4%) |
| Regional lymph node metastasis (cN1) | 4484 (3·4%) | 3027 (3·4%) | 668 (3·2%) | 527 (4·2%) | 221 (3·2%) | 27 (4·8%) |
| **Distant metastasis** |  |  |  |  |  |  |
| M1a, distant lymph nodes | 313 (0·2%) | 210 (0·2%) | 58 (0·3%) | 27 (0·2%) | 13 (0·2%) | 3 (0·5%) |
| M1b, bone | 3450 (2·6%) | 2253 (2·6%) | 569 (2·7%) | 392 (3·1%) | 208 (3·0%) | 22 (3·9%) |
| M1c, other sites | 715 (0·5%) | 421 (0·5%) | 147 (0·7%) | 97 (0·8%) | 42 (0·6%) | 7 (1·2%) |
| **NCCN risk group** |  |  |  |  |  |  |
| Low | 44 076 (33·7%) | 30 661 (34·8%) | 6296 (30·0%) | 4397 (34·7%) | 2005 (28·7%) | 171 (30·1%) |
| Intermediate | 52 736 (40·4%) | 35 847 (40·6%) | 8782 (41·9%) | 4760 (37·6%) | 2702 (38·7%) | 198 (34·9%) |
| High, very high, N1 or M1 | 33 822 (25·9%) | 21 718 (24·6%) | 5890 (28·1%) | 3517 (27·7%) | 2270 (32·5%) | 199 (35·0%) |
| **Marital status** |  |  |  |  |  |  |
| Married | 86 767 (66·4%) | 61 637 (69·9%) | 10 976 (52·3%) | 8157 (64·4%) | 5061 (72·5%) | 340 (59·9%) |
| Single | 13 996 (10·7%) | 7821 (8·9%) | 4088 (19·5%) | 1389 (11·0%) | 528 (7·6%) | 66 (11·6%) |
| Divorced/Separated | 11 073 (8·5%) | 6861 (7·8%) | 2753 (13·1%) | 1041 (8·2%) | 298 (4·3%) | 73 (12·9%) |
| Widowed | 4318 (3·3%) | 2953 (3·3%) | 763 (3·6%) | 383 (3·0%) | 185 (2·7%) | 18 (3·2%) |
| Unknown | 14 480 (11·1%) | 8954 (10·1%) | 2388 (11·4%) | 1704 (13·4%) | 905 (13·0%) | 71 (12·5%) |
| **County median household income** |  |  |  |  |  |  |
| Q1 (lowest) | 35 077 (26·9%) | 24 273 (27·5%) | 7824 (37·3%) | 2312 (18·2%) | 365 (5·2%) | 191 (33·6%) |
| Q2 | 32 216 (24·7%) | 18 910 (21·4%) | 6343 (30·3%) | 4884 (38·5%) | 1571 (22·5%) | 134 (23·6%) |
| Q3 | 34 240 (26·2%) | 24 967 (28·3%) | 4081 (19·5%) | 2734 (21·6%) | 1978 (28·4%) | 182 (32·0%) |
| Q4 (highest) | 29 101 (22·3%) | 20 076 (22·8%) | 2720 (13·0%) | 2744 (21·7%) | 3063 (43·9%) | 61 (10·7%) |
| **Residence** |  |  |  |  |  |  |
| Metro ≥1M | 76 919 (58·9%) | 48 776 (55·3%) | 13 399 (63·9%) | 8694 (68·6%) | 4936 (70·7%) | 221 (38·9%) |
| Metro 250k-1M | 28 737 (22·0%) | 19 641 (22·3%) | 4116 (19·6%) | 2823 (22·3%) | 1776 (25·5%) | 119 (21·0%) |
| Metro <250k | 10 234 (7·8%) | 7657 (8·7%) | 1707 (8·1%) | 666 (5·3%) | 111 (1·6%) | 70 (12·3%) |
| Nonmetro adjacent | 8644 (6·6%) | 7030 (8·0%) | 1250 (6·0%) | 259 (2·0%) | 23 (0·3%) | 53 (9·3%) |
| Nonmetro nonadjacent | 6027 (4·6%) | 5106 (5·8%) | 496 (2·4%) | 228 (1·8%) | 130 (1·9%) | 53 (9·3%) |
| **Initial treatment (registry-recorded)** |  |  |  |  |  |  |
| Radical prostatectomy | 46 706 (35·8%) | 33 307 (37·8%) | 5979 (28·5%) | 4521 (35·7%) | 2485 (35·6%) | 178 (31·3%) |
| Radiotherapy | 50 402 (38·6%) | 33 017 (37·4%) | 9187 (43·8%) | 4783 (37·7%) | 2749 (39·4%) | 211 (37·1%) |
| Chemotherapy | 1051 (0·8%) | 720 (0·8%) | 166 (0·8%) | 99 (0·8%) | 54 (0·8%) | 6 (1·1%) |
| **Deaths during follow-up** |  |  |  |  |  |  |
| Prostate cancer | 9575 (7·3%) | 6436 (7·3%) | 1649 (7·9%) | 958 (7·6%) | 448 (6·4%) | 74 (13·0%) |
| Other causes | 24 952 (19·1%) | 17 120 (19·4%) | 4485 (21·4%) | 2035 (16·1%) | 1122 (16·1%) | 133 (23·4%) |

Data are n (%) or median (IQR). Percentages may not sum to 100 because of rounding. Race and ethnicity unknown (n=1221) included in the overall column only. NCCN=National Comprehensive Cancer Network (see Methods). Treatment as recorded by registries (known to be under-ascertained).

Discrimination was high and stable across horizons (figure 2; table 2). At 10 years, XGBoost achieved a time-dependent AUC of 0·872 (0·867–0·877) for PCSM, 0·742 (0·739–0·746) for OCM and 0·795 (0·792–0·799) for all-cause mortality. The spline-based regression model performed almost identically (0·869 [0·864–0·873], 0·741 [0·738–0·745] and 0·794 [0·791–0·797]). The difference in AUC between XGBoost and regression was statistically detectable for every outcome and horizon but never exceeded 0·005. Across all five models, the 10-year AUC ranges were 0·004 (PCSM), 0·005 (OCM) and 0·003 (all-cause). All models outperformed the clinical risk tools. For PCSM, the best-performing tool was UCSF-CAPRA, with an AUC of 0·838, a difference of 0·034 (0·032–0·036) relative to XGBoost. The gap was larger for all-cause mortality (difference 0·087 [0·084–0·091]) and for OCM (difference 0·128 [0·124–0·133]; tool AUCs ≤0·61), as expected for tools that were not designed to predict death from other causes.

**Figure 2:**
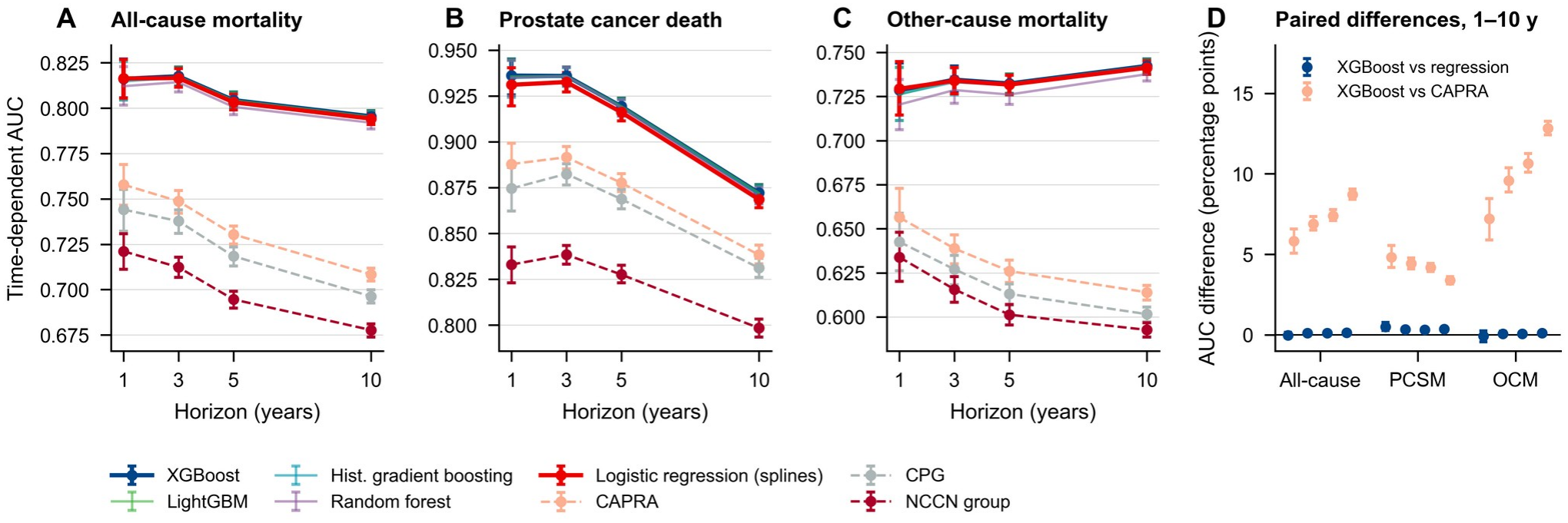
Discrimination across prediction horizons. Time-dependent AUC (competing-risk definition, inverse probability of censoring weighted) of cross-validated predictions for (A) all-cause mortality, (B) prostate cancer-specific mortality and (C) other-cause mortality at 1, 3, 5 and 10 years, for the five competing-risk models (solid lines) and risk-stratification tools (dashed lines). (D) Paired bootstrap differences in AUC (percentage points) between XGBoost and logistic regression (blue) and between XGBoost and CAPRA (orange) at 1, 3, 5 and 10 years. Error bars are 95% CIs.

**Table 2:**
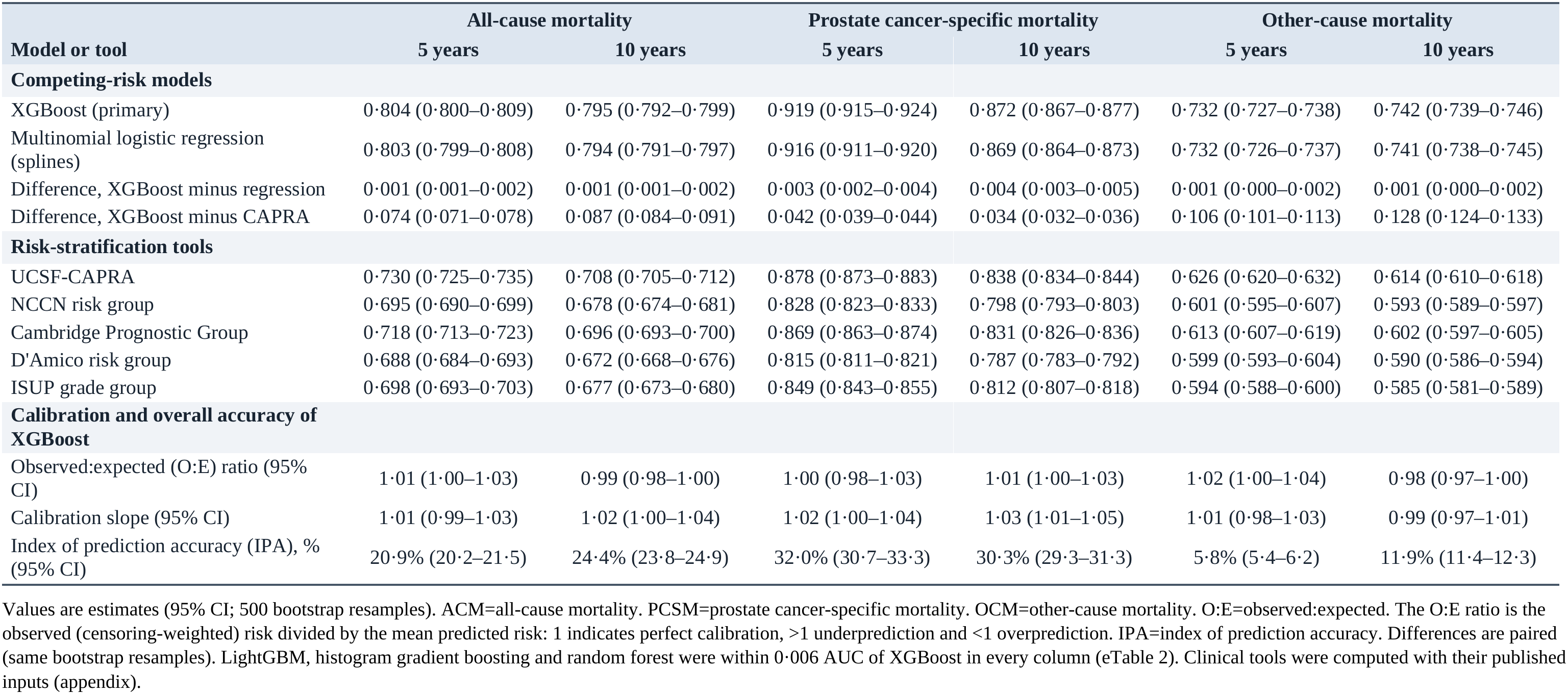
Discrimination (time-dependent AUC) and calibration of cross-validated predictions at 5 and 10 years.

|  | All-cause mortality |  | Prostate cancer-specific mortality |  | Other-cause mortality |  |
|---|---|---|---|---|---|---|
| **Model or tool** | **5 years** | **10 years** | **5 years** | **10 years** | **5 years** | **10 years** |
| **Competing-risk models** |  |  |  |  |  |  |
| XGBoost (primary) | 0·804 (0·800–0·809) | 0·795 (0·792–0·799) | 0·919 (0·915–0·924) | 0·872 (0·867–0·877) | 0·732 (0·727–0·738) | 0·742 (0·739–0·746) |
| Multinomial logistic regression (splines) | 0·803 (0·799–0·808) | 0·794 (0·791–0·797) | 0·916 (0·911–0·920) | 0·869 (0·864–0·873) | 0·732 (0·726–0·737) | 0·741 (0·738–0·745) |
| Difference, XGBoost minus regression | 0·001 (0·001–0·002) | 0·001 (0·001–0·002) | 0·003 (0·002–0·004) | 0·004 (0·003–0·005) | 0·001 (0·000–0·002) | 0·001 (0·000–0·002) |
| Difference, XGBoost minus CAPRA | 0·074 (0·071–0·078) | 0·087 (0·084–0·091) | 0·042 (0·039–0·044) | 0·034 (0·032–0·036) | 0·106 (0·101–0·113) | 0·128 (0·124–0·133) |
| **Risk-stratification tools** |  |  |  |  |  |  |
| UCSF-CAPRA | 0·730 (0·725–0·735) | 0·708 (0·705–0·712) | 0·878 (0·873–0·883) | 0·838 (0·834–0·844) | 0·626 (0·620–0·632) | 0·614 (0·610–0·618) |
| NCCN risk group | 0·695 (0·690–0·699) | 0·678 (0·674–0·681) | 0·828 (0·823–0·833) | 0·798 (0·793–0·803) | 0·601 (0·595–0·607) | 0·593 (0·589–0·597) |
| Cambridge Prognostic Group | 0·718 (0·713–0·723) | 0·696 (0·693–0·700) | 0·869 (0·863–0·874) | 0·831 (0·826–0·836) | 0·613 (0·607–0·619) | 0·602 (0·597–0·605) |
| D'Amico risk group | 0·688 (0·684–0·693) | 0·672 (0·668–0·676) | 0·815 (0·811–0·821) | 0·787 (0·783–0·792) | 0·599 (0·593–0·604) | 0·590 (0·586–0·594) |
| ISUP grade group | 0·698 (0·693–0·703) | 0·677 (0·673–0·680) | 0·849 (0·843–0·855) | 0·812 (0·807–0·818) | 0·594 (0·588–0·600) | 0·585 (0·581–0·589) |
| **Calibration and overall accuracy of XGBoost** |  |  |  |  |  |  |
| Observed:expected (O:E) ratio (95% CI) | 1·01 (1·00–1·03) | 0·99 (0·98–1·00) | 1·00 (0·98–1·03) | 1·01 (1·00–1·03) | 1·02 (1·00–1·04) | 0·98 (0·97–1·00) |
| Calibration slope (95% CI) | 1·01 (0·99–1·03) | 1·02 (1·00–1·04) | 1·02 (1·00–1·04) | 1·03 (1·01–1·05) | 1·01 (0·98–1·03) | 0·99 (0·97–1·01) |
| Index of prediction accuracy (IPA), % (95% CI) | 20·9% (20·2–21·5) | 24·4% (23·8–24·9) | 32·0% (30·7–33·3) | 30·3% (29·3–31·3) | 5·8% (5·4–6·2) | 11·9% (11·4–12·3) |

Values are estimates (95% CI; 500 bootstrap resamples). ACM=all-cause mortality. PCSM=prostate cancer-specific mortality. OCM=other-cause mortality. O:E=observed:expected. The O:E ratio is the observed (censoring-weighted) risk divided by the mean predicted risk: 1 indicates perfect calibration, >1 underprediction and <1 overprediction. IPA=index of prediction accuracy. Differences are paired (same bootstrap resamples). LightGBM, histogram gradient boosting and random forest were within 0·006 AUC of XGBoost in every column (eTable 2). Clinical tools were computed with their published inputs (appendix).

XGBoost and regression were well calibrated across the full range of predicted risk (figure 3). At 10 years, XGBoost had O:E ratios of 1·01, 0·98 and 0·99 for PCSM, OCM and all-cause mortality, calibration slopes of 1·03, 0·99 and 1·02, and ICIs of 0·24, 0·35 and 0·37 percentage points. Regression gave similar values (slopes 1·07, 0·99 and 1·05). The random forest was systematically miscalibrated (10-year slopes 1·12–1·15), suggesting that its predictions were too close to the average risk (appendix). IPA values for XGBoost were 30·3% (PCSM), 11·9% (OCM) and 24·4% (all-cause) relative to a null model.

**Figure 3:**
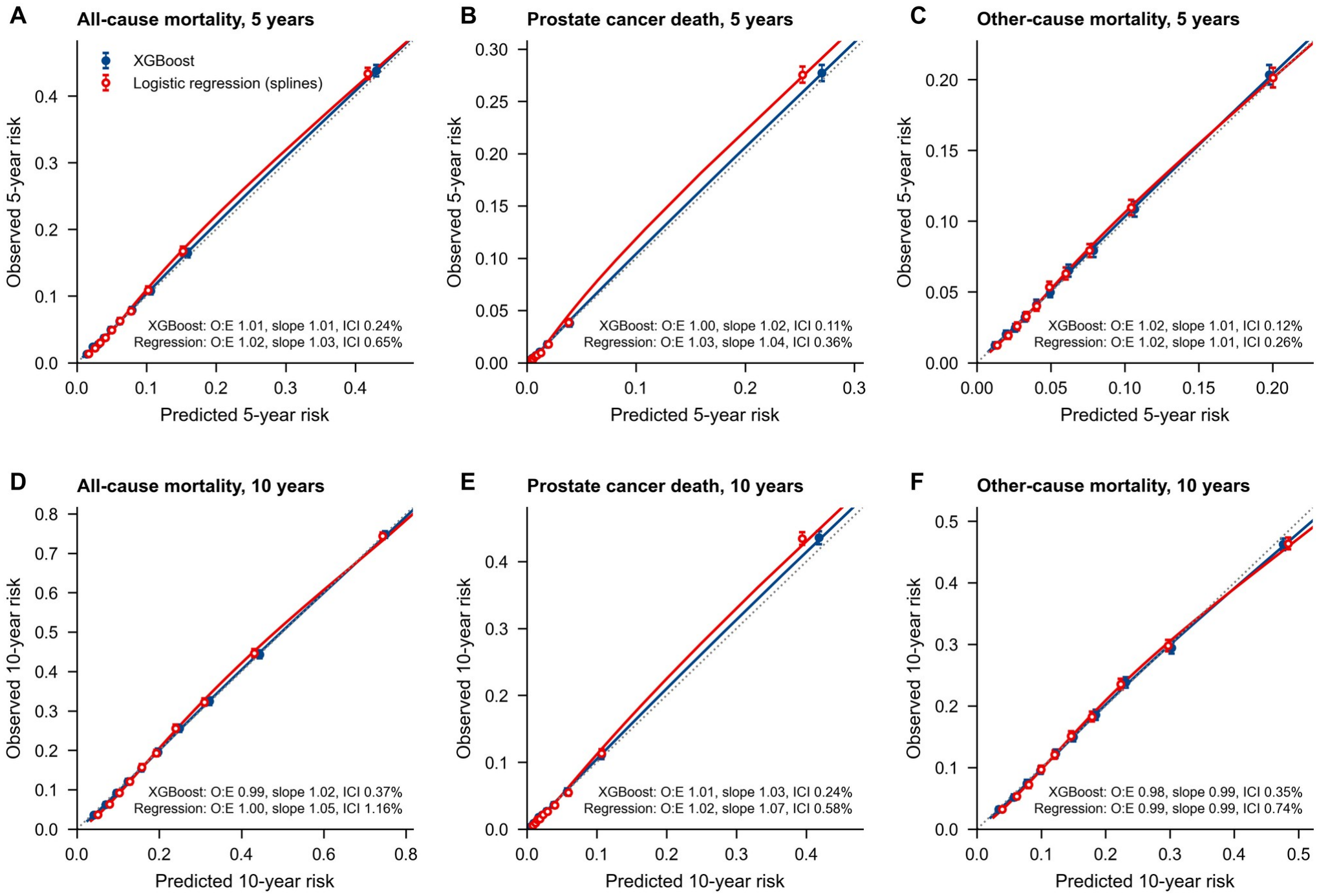
Calibration of predicted 5-year and 10-year risks. Points show mean predicted versus inverse probability of censoring-weighted observed risk in deciles of predicted risk (95% CI). Lines are flexible calibration curves (restricted cubic splines). The dotted line indicates perfect calibration. O:E=observed:expected ratio. ICI=integrated calibration index.

In the temporal split (training on 2010–2012 diagnoses and evaluation on 2013–2015 diagnoses), 5- year AUCs for XGBoost were 0·923 (0·918–0·929) for PCSM, 0·726 (0·719–0·734) for OCM and 0·807 (0·802–0·813) for all-cause mortality (regression 0·921, 0·728 and 0·808), with O:E ratios of 0·96, 0·94 and 0·95. The models slightly overpredicted mortality in the later cohort, suggesting that calibration may need to be reassessed over time (appendix).

In decision-curve analysis (appendix), the competing-risk models gave the highest net benefit across clinically relevant thresholds for all three outcomes. For 10-year PCSM at a 10% threshold, net benefit was 0·0367 for XGBoost versus 0·0308 for CAPRA and −0·0340 for treating all men as high risk. For 10-year all-cause mortality at a 30% threshold, net benefit was 0·0852 versus 0·0554 for CAPRA. Regression and XGBoost curves were nearly indistinguishable, with a marginal advantage for XGBoost only at PCSM thresholds above 20%.

PCSM and all-cause mortality predictions performed similarly across racial and ethnic groups (figure 4; table 3). At 10 years, XGBoost AUCs ranged from 0·852 to 0·887 for PCSM and from 0·762 to 0·802 for all-cause mortality. O:E ratios ranged from 1·01 to 1·09 for PCSM and from 0·98 to 1·05 for all-cause mortality. CIs were wide for American Indian or Alaska Native men (n=568). Discrimination for OCM was lower in Black men (0·708) and American Indian or Alaska Native men (0·679) than in White men (0·746). Excluding race and ethnicity from the model barely changed discrimination within groups (differences −0·001 to 0·001) but degraded calibration for non-Hispanic Black men. The model excluding race and ethnicity underpredicted their 10-year OCM (O:E 1·16 [1·13–1·19], versus 0·99 with race and ethnicity included) and all-cause mortality (O:E 1·12 [1·09– 1·14]), with PCSM O:E 1·02 (0·98–1·06). Conversely, it overpredicted OCM in Asian or Pacific Islander men (O:E 0·80 [0·76–0·85]) and Hispanic men (0·85 [0·81–0·89]). The same pattern was seen with the regression model excluding race and ethnicity (appendix) and for all-cause mortality, which does not depend on cause-of-death attribution.

**Figure 4:**
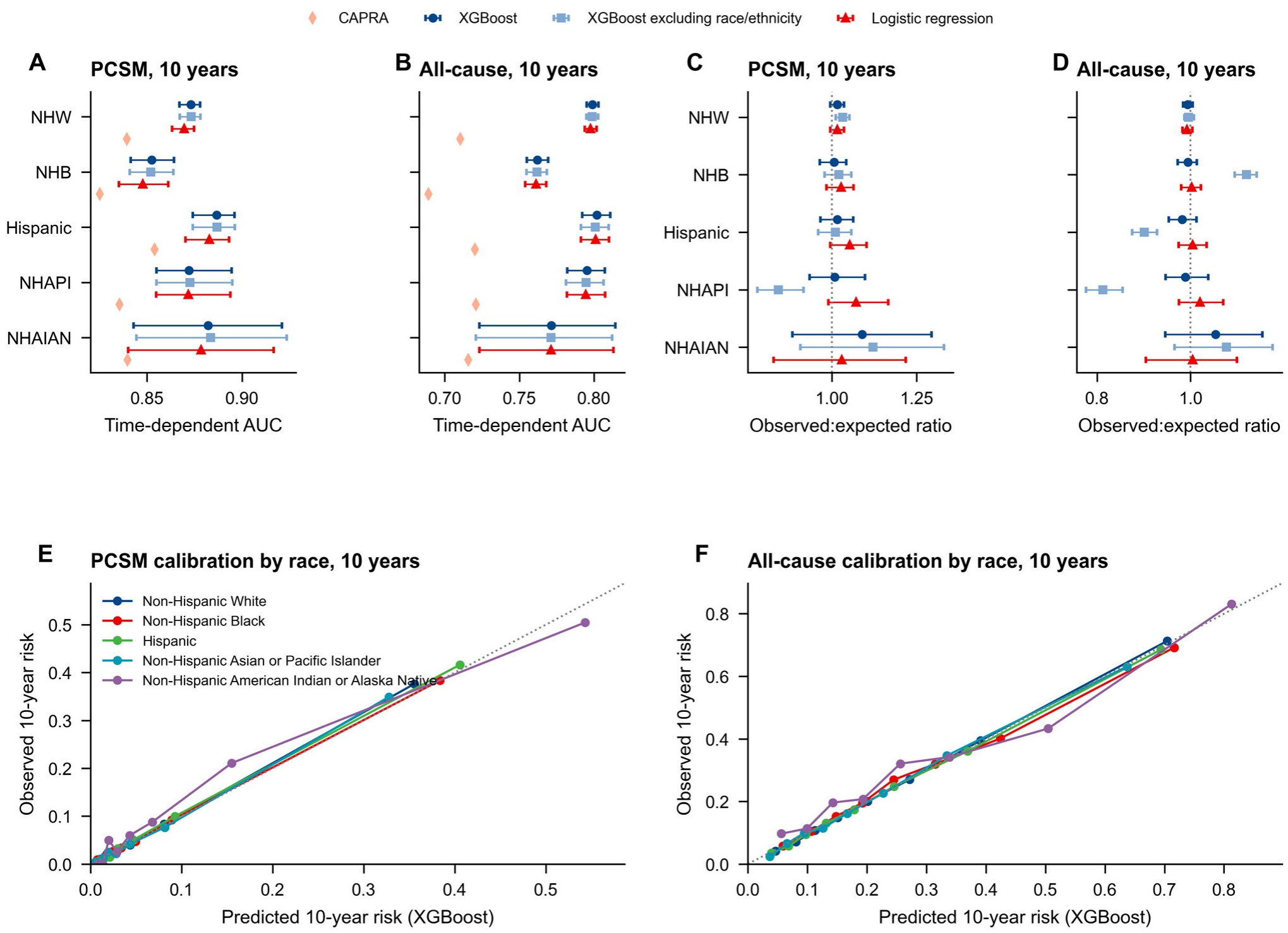
Race-stratified validation at 10 years. (A, B) Time-dependent AUC and (C, D) observed:expected ratios (95% CI) for prostate cancer-specific and all-cause mortality in each racial and ethnic group for XGBoost, XGBoost excluding race and ethnicity, and logistic regression. Diamonds show CAPRA. (E, F) Calibration of XGBoost by racial and ethnic group (deciles of predicted risk). NHW=non-Hispanic White. NHB=non-Hispanic Black. NHAPI=non-Hispanic Asian or Pacific Islander. NHAIAN=non-Hispanic American Indian or Alaska Native.

**Table 3:** Race-stratified performance of XGBoost for 10-year outcomes.

| Race and ethnicity | n | Prostate cancer-specific mortality |  | All-cause mortality |  | Other-cause mortality |  |  |
|---|---|---|---|---|---|---|---|---|
|  |  | AUC | O:E ratio | AUC | O:E ratio | O:E ratio, including race and ethnicity | O:E ratio, excluding race and ethnicity | Reclassified, % |
| Non-Hispanic White | 88 226 | 0·873 (0·867–0·878) | 1·02 (1·00–1·04) | 0·799 (0·795–0·803) | 0·99 (0·98–1·00) | 0·98 (0·97–1·00) | 0·98 (0·97–0·99) | 0·6 |
| Non-Hispanic Black | 20 968 | 0·852 (0·841–0·864) | 1·01 (0·96–1·04) | 0·762 (0·755–0·769) | 0·99 (0·97–1·01) | 0·99 (0·96–1·02) | 1·16 (1·13–1·19) | 2·1 |
| Hispanic | 12 674 | 0·887 (0·874–0·896) | 1·02 (0·97–1·06) | 0·802 (0·792–0·811) | 0·98 (0·95–1·01) | 0·97 (0·92–1·00) | 0·85 (0·81–0·89) | 1·6 |
| Non-Hispanic Asian or Pacific Islander | 6977 | 0·872 (0·855–0·894) | 1·01 (0·93–1·10) | 0·795 (0·782–0·807) | 0·99 (0·95–1·04) | 0·98 (0·93–1·04) | 0·80 (0·76–0·85) | 3·8 |
| Non-Hispanic American Indian or Alaska Native | 568 | 0·882 (0·843–0·921) | 1·09 (0·88–1·29) | 0·771 (0·723–0·814) | 1·05 (0·95–1·15) | 1·03 (0·86–1·22) | 1·05 (0·88–1·24) | 1·6 |

Values are estimates (95% CI from bootstrap resampling within each group). AUC=time-dependent area under the receiver operating characteristic curve. O:E=observed:expected ratio (observed risk divided by mean predicted risk). O:E>1 indicates underprediction and <1 overprediction. The model excluding race and ethnicity is XGBoost refitted without these variables. Reclassified: classification relative to a 10-year life expectancy (predicted 10-year all-cause mortality ≥50%) differs between the models including and excluding race and ethnicity.

These calibration differences changed which men crossed the prespecified threshold (figure 5). At the 10-year life-expectancy threshold, the model excluding race and ethnicity reclassified 2·1% of Black men (3·0% of those with non-metastatic NCCN intermediate or high-risk disease), almost all toward a longer life expectancy, and 3·8% of Asian or Pacific Islander men (5·1%) and 1·6% of Hispanic men toward a shorter one, compared with 0·6% of White men. Observed 10-year mortality among the reclassified men was 49·2% in Black men (95% CI 43·7–53·9, n=440), 40·8% in Asian or Pacific Islander men (95% CI 33·7–47·4, n=267) and 53·4% in Hispanic men (95% CI 45·0–62·1, n=190).

**Figure 5:**
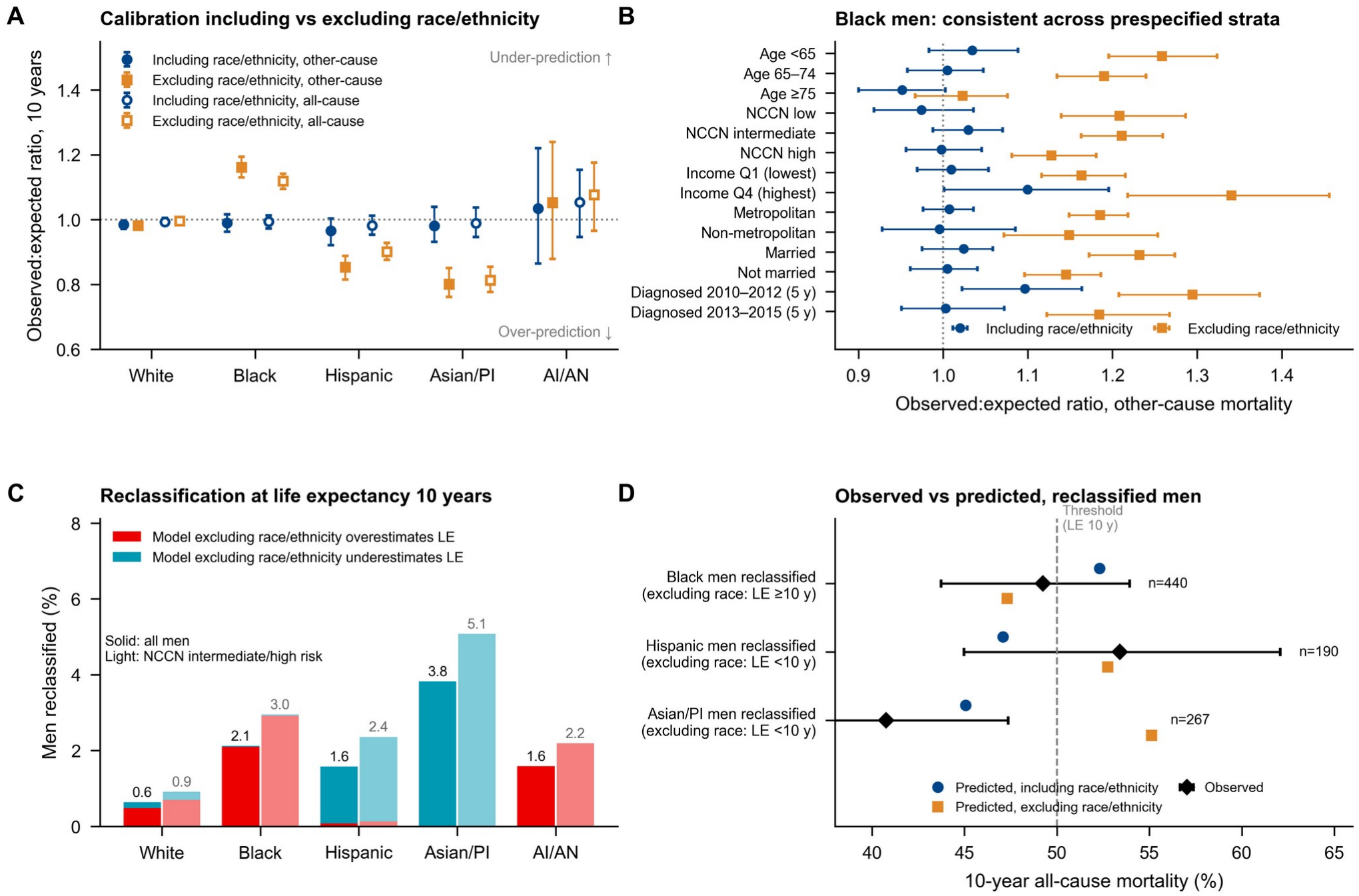
Consequences of excluding race and ethnicity from the model. (A) Observed:expected ratios (95% CI) for 10-year other-cause (filled) and all-cause (open) mortality with the XGBoost model including race and ethnicity (blue) and the model excluding race and ethnicity (orange). (B) Other-cause mortality O:E in non-Hispanic Black men across prespecified strata (10 years; diagnosis period at 5 years). (C) Percentage of men whose classification relative to a 10-year life expectancy (predicted 10-year all-cause mortality ≥50%) differs between the two models, for all men (solid) and men with non-metastatic NCCN intermediate or high-risk disease (light). (D) Observed 10-year all-cause mortality (95% CI) of reclassified men compared with the mean risk predicted by each model. The dashed line is the life-expectancy threshold. AI/AN=American Indian or Alaska Native. PI=Pacific Islander. LE=life expectancy.

For these groups, the corresponding predictions were 52·3%, 45·1% and 47·1% with race and ethnicity included, and 47·3%, 55·1% and 52·8% with them excluded. In Asian or Pacific Islander men, the confidence interval excluded the prediction of the model excluding race and ethnicity but not that of the model including them. In Black and Hispanic men, observed mortality was close to the threshold and the intervals included both predictions. In Black men, underprediction of OCM by the model excluding race and ethnicity was present in every prespecified stratum (O:E 1·02–1·34, versus 0·95–1·10 with race), including the highest income quartile (1·34), and was largest in men younger than 65 years (1·26). Because reclassification affected a minority of men close to the threshold, population-level differences in net benefit were small (Black men −0·5 per 1000, 95% CI −2·7 to 1·8).

The 10-year cumulative incidence of PCSM was 7·5% in non-Hispanic Black men and 6·9% in non-Hispanic White men; for OCM it was 19·6% and 17·0% (appendix). Relative to non-Hispanic White men, the age-adjusted cause-specific hazard ratio for PCSM in non-Hispanic Black men was 1·31 (1·24–1·38). After adjustment for tumor characteristics it was 1·09 (1·03–1·16). Confidence intervals included 1 after further adjustment for social factors (1·01 [0·95–1·07]) and treatment (0·96 [0·91– 1·02]). Excess OCM in non-Hispanic Black men (1·50 [1·45–1·55] age-adjusted) persisted after full adjustment (1·21 [1·17–1·25]). Asian or Pacific Islander men had lower PCSM and OCM in every model (0·66 [0·60–0·73] and 0·76 [0·71–0·81] fully adjusted). Lowest-quartile income and non-metropolitan residence were independently associated with higher OCM (1·44 [1·39–1·50] and 1·11 [1·07–1·16]) (appendix).

The features most important to the model differed by cause of death (appendix figure). Tumor characteristics (PSA, grade group, clinical T, N and M stage and biopsy-core involvement) accounted for 88% of the explained variation in the PCSM hazard but 16% for the OCM hazard, which was dominated by age (48%) and social factors (marital status 12%, income 16%, race and ethnicity 7%).

In sensitivity analyses, horizon-specific XGBoost classifiers that accounted for competing risks performed similarly to the single discrete-time model (10-year PCSM AUCs of 0·871 and 0·872, with O:E ratios of 1·00 and 1·01, respectively). Excluding men who died of other causes produced similar discrimination (0·869) but overestimated PCSM risk by 36% at 10 years (O:E 0·73) and 15% at 5 years (O:E 0·87). Adding first-course treatment changed the 10-year AUC by 0·005 (0·004–0·006) for PCSM and 0·008 (0·007–0·009) for all-cause mortality (appendix).

Compared with the registry-minimal model (age, PSA, Gleason score and social variables) in the same men, the clinical model improved the 10-year AUC by 0·015 (0·014–0·017) for PCSM, 0·006 (0·006– 0·007) for all-cause mortality and 0·002 (0·002–0·003) for OCM. The calibration shift after excluding race and ethnicity was similar in the registry-minimal model (10-year OCM O:E in Black men 0·99 [0·96–1·02] with race and 1·17 [1·14–1·20] without; Asian or Pacific Islander men 0·98 [0·93–1·03] and 0·80 [0·76–0·85], appendix).

Results were similar without the biopsy-core requirement (205 933 men with known clinical T, N and M categories, appendix). The clinical model without core counts had 10-year AUCs of 0·873 (0·870– 0·876) for PCSM, 0·746 (0·743–0·748) for OCM and 0·798 (0·796–0·801) for all-cause mortality, and excluding race and ethnicity again shifted OCM calibration (Black men 0·99 [0·97–1·01] with race and 1·18 [1·15–1·20] without; Asian or Pacific Islander men 0·98 [0·93–1·03] and 0·79 [0·75– 0·83]). Within the men of the primary cohort, core counts improved the 10-year AUC by 0·002 (0·002 –0·004) for PCSM and 0·001 (0·001–0·002) for all-cause mortality.

## Discussion

In 130 634 men with prostate adenocarcinoma, one discrete-time competing-risk model predicted prostate cancer-specific, other-cause and all-cause mortality from 1 to 10 years after diagnosis, with high discrimination and good calibration overall, in a temporal split and across racial and ethnic groups. Machine learning and flexible regression performed similarly (maximum AUC difference 0·005, IPA difference ≤1·0 percentage points), consistent with systematic evidence that machine learning rarely outperforms regression when predictors are few and well measured.^20^ Earlier SEER studies compared algorithms with categorical risk groups or standard Cox models.^18,19^ The regression model can be published as a formula and is the basis of the web calculator.

Tools that stratify cancer risk had lower discrimination for other-cause and all-cause mortality. Guidelines consider both outcomes in treatment decisions.^7–9^ The choice between surveillance and radical treatment depends on the balance between these risks.^5,6^ PREDICT Prostate already estimates both outcomes.^16^ We estimated these risks over several time horizons in a racially diverse cohort. Ignoring competing deaths when labeling outcomes, as many machine-learning studies do, had little effect on discrimination but overestimated 10-year prostate cancer mortality by 36%. In decision-curve analysis, the net benefit of XGBoost exceeded that of CAPRA by 0·0059 at a 10% threshold for 10- year prostate cancer death, equivalent in net benefit to identifying about six more men per 1000 at the same false-positive rate. The difference in net benefit was 0·0298 (about 30 per 1000) at a 30% threshold for 10-year all-cause death. Regression performed similarly.

Fairness was examined as TRIPOD+AI recommends.^28,31^ Excluding race and ethnicity left within-group discrimination unchanged but underpredicted other-cause mortality in Black men and overpredicted it in Asian or Pacific Islander and Hispanic men, moving up to one in 20 men with non-metastatic intermediate or high-risk disease across the study’s life-expectancy threshold. Among reclassified men, observed mortality favored the model including race and ethnicity only in the Asian or Pacific Islander subgroup. Underprediction in Black men persisted in the highest income quartile and was greater in men younger than 65 years. The pattern was similar in models with and without detailed tumor characteristics. These differences may reflect health and social factors that the registry does not capture.^26,27^ Discrimination metrics did not detect the differences that we observed in subgroup calibration. Similar miscalibration followed the exclusion of race in colorectal cancer recurrence models^45^; race-neutral life tables endorsed for estimating life expectancy underestimate mortality in Black men with prostate cancer^46^; and an other-cause mortality model that measured comorbidity directly performed well without race.^47^ Race and ethnicity therefore appear to carry information about competing mortality that is not otherwise measured. Future studies should measure factors such as comorbidity and frailty directly and evaluate calibration by race and ethnicity.^25,48^

Differences in tumor characteristics at diagnosis accounted for much of the higher prostate cancer mortality in Black men, consistent with similar outcomes under equal access to care.^29,30^ Their excess other-cause mortality persisted after adjustment for tumor, social and treatment factors, which may reflect differences in comorbidity and social factors that the registry does not measure.

Our study has limitations. SEER lacks MRI findings, genomic classifiers, comorbidity, performance status and, since the November 2019 release, insurance status. Registry treatment data are incomplete,^49^ and cause-of-death attribution from death certificates is imperfect, although the SEER algorithm reduces misclassification.^34^ The complete-case restriction excluded men who were older and had higher mortality, mostly because biopsy-core counts were not recorded, but the main findings were similar when we relaxed this requirement. The diagnoses predate routine MRI-targeted biopsy. Validation, including the temporal split, was internal to SEER. External validation and prospective evaluation are required before implementation.^50,51^ The mortality threshold used to approximate life expectancy differs from guideline-based life expectancy estimates.

Using clinical data available at diagnosis, the models estimated both prostate cancer and other-cause mortality with good calibration. Flexible regression and machine learning performed similarly. Excluding competing deaths overestimated prostate cancer mortality. Studies of prognostic tools should assess both outcomes and report calibration across racial and ethnic groups.

## Data Availability

All data produced in the present study are available upon reasonable request to the authors.

https://seer.cancer.gov/data/access.html

## Declaration of interests

The authors declare no competing interests.

## Data sharing

SEER data are available from the National Cancer Institute under a data-use agreement (https://seer.cancer.gov/data/access.html). The analysis code, the fitted model files and the prediction formula of the regression model will be made publicly available on publication, together with an interactive web calculator that reproduces the regression model’s predictions exactly (research version, https://immuneresponse.github.io/prostate-risk-calculator/).

## Acknowledgments

We thank the SEER registries and the Surveillance Research Program of the National Cancer Institute. The interpretation and reporting of these data are the sole responsibility of the authors. OpenAI Codex was used for language editing and manuscript formatting.

## Notes

### Competing Interest Statement

The authors have declared no competing interest.

### Author Declarations

Secondary analysis of de-identified SEER Research Data accessed through NCI registration and a data-use agreement. There was no contact with participants. An approval or waiver from an institutional ethics oversight body has not been supplied for this submission.

