## Supplementary Appendix for "Population-based multi-horizon machine-learning prognostication for prostate adenocarcinoma: 10-year competing-risk and race-stratified validation"

eMethods 1. Data audit of the SEER extract
eMethods 2. Variable definitions and risk-stratification tools
eMethods 3. Discrete-time competing-risk model and performance measures
eMethods 4. Verification of the metric code by simulation
eTable 1. Characteristics of included and excluded men
eTable 2. Performance of all models and tools at all horizons
eTable 3. Temporal validation
eTable 4. Race-stratified performance, all outcomes and horizons
eTable 5. Race-aware versus race-blind models
eTable 6. Treatment-augmented and registry-minimal models
eTable 7. Sensitivity analysis: direct and naive horizon-specific classifiers
eTable 8. Cause-specific hazard ratios for race and ethnicity (sequential adjustment)
eTable 9. Cumulative incidence by race and ethnicity and by NCCN group
eFigure 1. Competing-mortality landscape and model explanation (SHAP)
eFigure 2. Cumulative incidence by race and ethnicity
eFigure 3. Temporal validation
eFigure 4. Calibration of all models at all horizons
eFigure 5. Decision-curve analysis
eTable 10. Race-blind calibration shift across prespecified strata
eMethods 5 and eTables 11–12. Temporal validation in men diagnosed in 2016–2017
eFigure 6. Calibration in men diagnosed in 2016–2017
eMethods 6 and eTable 13. Fixed 10-year binary classification in a held-out test set
eFigures 7–9. ROC curves, risk-group cumulative incidence, and SHAP
TRIPOD+AI checklist

### eMethods 1. Data audit of the SEER extract

Data came from two unfiltered SEER*Stat case listings of the same database, each containing 9 708 868 tumour records of all sites in the same order, exported as text. The first held survival, cause of death, tumour and demographic fields (1 334 542 prostate records); the second held sequence number, county income and rurality, first-course treatment, single-year age and derived T stage for 2016–2017. The exports were joined on record position. Because a patient can have several primaries diagnosed in one year, patient identifier and year of diagnosis do not identify a tumour uniquely, so the join was accepted only after confirming that both fields agreed on every record (0 disagreements). All prostate records were linked. An earlier spreadsheet extract used for the conference abstract was truncated at the spreadsheet row limit and is not used here. All analyses are fully scripted.

### eMethods 2. Variable definitions and risk-stratification tools

Age: single year of age at diagnosis (90+ coded as 92). PSA: PSA Lab Value Recode (2010+), in ng/mL; the category '98·0 ng/mL or greater' was set to 98. Gleason score: Gleason Score Clinical Recode (2010+) (biopsy or transurethral resection). Grade group: from the Gleason Patterns Clinical Recode (3+3 or less: 1; 3+4: 2; 4+3: 3; sum 8: 4; sum 9–10: 5); when the patterns were unknown, from the Gleason sum (7 assigned grade group 2). Clinical T category: CS Extension – Clinical Extension (codes 100–199 T1; 210 T2a, half of one lobe or less; 220 T2b, more than half of one lobe; 230 T2c, both lobes; 200, 240 and 300 T2 not otherwise specified; 400–499 T3a; 500–599 T3b; 600–799 T4; 000, 800, 950 and 999 unknown). Regional nodes: CS Lymph Nodes (000 cN0; 100–800 cN1; 999 unknown). Distant metastasis: CS Mets at DX (00 M0; 10–12 M1a, distant lymph nodes; 20–38 M1b, bone; 40–60 M1c, other sites; 99 unknown). Biopsy cores: CS site-specific factors 12 (positive) and 13 (examined); usable when both were recorded counts (0–99 positive, 1–99 examined, positive ≤ examined); codes for unknown or no needle biopsy were treated as missing. Income: county median household income, inflation-adjusted to 2024, band midpoints and cohort quartiles. Rurality: Rural–Urban Continuum Code, five categories. Radical prostatectomy: surgery of primary site codes 50, 70 or 80. Radiotherapy: beam, implants, radioisotopes, combination or not otherwise specified. Chemotherapy: chemotherapy recode 'Yes'.

UCSF-CAPRA (0–10): age ≥50 years 1 point; PSA ≤6 0, 6·01–10 1, 10·01–20 2, 20·01–30 3, >30 4 points; Gleason: no pattern 4 or 5 0, secondary pattern 4 or 5 1 (grade group 2), primary pattern 4 or 5 3 points (grade group 3–5); clinical T3a or higher 1 point; ≥34% positive cores 1 point.

NCCN risk group: low (T1–T2a, grade group 1 and PSA <10), intermediate (T2b–T2c, grade group 2–3 or PSA 10–20), high (T3–T4, grade group 4–5 or PSA >20, including very high risk, cN1 and M1). T2 not otherwise specified was treated as T2a. The very-low and favourable or unfavourable intermediate subdivisions were not separated.

Cambridge Prognostic Group: CPG1 grade group 1, PSA <10 and T1–T2; CPG2 grade group 2 or PSA 10–20, and T1–T2; CPG3 grade group 3, or grade group 2 with PSA 10–20, and T1–T2; CPG4 exactly one of grade group 4, PSA >20 or T3; CPG5 two or more of these, or grade group 5, or T4.

D'Amico: low (T1–T2a, Gleason ≤6 and PSA ≤10), intermediate (T2b, Gleason 7 or PSA 10–20), high (T2c or higher, Gleason ≥8 or PSA >20).

### eMethods 3. Discrete-time competing-risk model and performance measures

Person-period expansion. For patient i with follow-up $T_i$ (completed intervals) and status $\delta_i \in \{0,1,2\}$, records $j=0,\ldots,\min(T_i,10)$ were created with outcome $y_{ij}=\delta_i$ if $j=T_i$ and 0 otherwise. The multinomial classifier estimates $\pi_k(x,j)=P(y=k|x,j)$, which equals the cause-specific discrete hazard $\lambda_k(j|x)$. Cumulative incidence was computed as in the main text; all-cause risk is $CIF_1+CIF_2 = 1-S(h)$.

Censoring weights. $G(s)=P(C\geq s)$ was estimated by Kaplan–Meier with censoring as the event, events preceding censoring at tied times. At horizon h, men with an event at $T\leq h$ received weight $1/G(T)$, men event-free through h received $1/G(h)$, and men censored before h received 0.

Time-dependent AUC (competing-risk definition 2 of Blanche et al.): cases are men with the event of interest by h; controls are all other men with known status, including those with the competing event. AUC = $\Sigma\Sigma\, w_i w_j[1(r_i>r_j)+½1(r_i=r_j)] / (\Sigma w_i \Sigma w_j)$.

Brier score = $\Sigma w_i(D_i-r_i)^2/\Sigma w_i$; IPA = 1 − Brier/Brier(null), where the null model predicts the marginal cumulative incidence. Calibration intercept: weighted logistic regression of D on an offset logit(r); slope: coefficient of logit(r). Flexible calibration: weighted logistic regression on a restricted cubic spline of logit(r) (knots at the 5th, 35th, 65th and 95th percentiles); ICI and E90 are the weighted mean and 90th percentile of |smoothed observed − predicted| among men with evaluable status. O:E = $\Sigma w_i D_i / \Sigma w_i r_i$, so that observed and expected risks refer to the same weighted population even when censoring depends on diagnosis period. Net benefit at threshold p: $[\Sigma w_i D_i 1(r_i \geq p) - \Sigma w_i(1-D_i)1(r_i \geq p)\cdot p/(1-p)]/\Sigma w_i$.

Hyperparameter tuning (selected values): {"XGB": {"max_depth": 4, "min_child_weight": 100}, "LGBM": {"num_leaves": 15, "min_child_samples": 50}, "HGB": {"max_leaf_nodes": 15, "min_samples_leaf": 500}}.

### eMethods 4. Verification of the metric code by simulation

Without censoring, the weighted AUC and Brier score reproduced scikit-learn exactly (including ties). In 200 000 simulated patients with discrete-time competing risks and independent random censoring, the censoring-weighted observed 5-year cumulative incidence (13·64%) matched the uncensored truth (13·68%); the AUC of the true risk was 0·768 with weighting versus 0·768 in the uncensored data. Calibration of the true risk gave slope 0·996, intercept −0·011 and O:E 0·991, and a two-fold over-prediction gave O:E 0·51 (script test_metrics.py).

**eTable 1. Characteristics of included men and men excluded for missing PSA or Gleason score**

| | n | Mean age, years | Non-Hispanic Black, % | Died, % | Died of prostate cancer, % |
|---|---|---|---|---|---|
| Excluded (missing PSA or Gleason) | 131 645 | 66·7 | 14·5 | 34·0 | 11·4 |
| Included | 130 634 | 64·9 | 16·1 | 26·4 | 7·3 |

**eTable 2a. Time-dependent AUC (95% CI) and calibration: all-cause mortality**

| | 1 year | 3 years | 5 years | 10 years |
|---|---|---|---|---|
| Multinomial logistic regression (splines) | 0·816 (0·806–0·827) | 0·817 (0·811–0·822) | 0·803 (0·799–0·808) | 0·794 (0·791–0·797) |
| Random forest | 0·812 (0·802–0·823) | 0·814 (0·809–0·819) | 0·801 (0·796–0·805) | 0·792 (0·789–0·795) |
| Histogram gradient boosting | 0·815 (0·805–0·826) | 0·817 (0·812–0·822) | 0·804 (0·800–0·808) | 0·795 (0·791–0·798) |
| XGBoost | 0·816 (0·806–0·827) | 0·818 (0·812–0·823) | 0·804 (0·800–0·809) | 0·795 (0·792–0·799) |
| LightGBM | 0·815 (0·805–0·826) | 0·817 (0·812–0·823) | 0·804 (0·800–0·809) | 0·795 (0·792–0·798) |
| UCSF-CAPRA | 0·758 (0·746–0·769) | 0·749 (0·742–0·755) | 0·730 (0·725–0·735) | 0·708 (0·705–0·712) |
| NCCN risk group | 0·721 (0·711–0·731) | 0·712 (0·707–0·718) | 0·695 (0·690–0·699) | 0·678 (0·674–0·681) |
| Cambridge Prognostic Group | 0·744 (0·732–0·755) | 0·738 (0·731–0·744) | 0·718 (0·713–0·723) | 0·696 (0·693–0·700) |
| D'Amico risk group | 0·714 (0·704–0·724) | 0·706 (0·700–0·712) | 0·688 (0·684–0·693) | 0·672 (0·668–0·676) |
| ISUP grade group | 0·719 (0·706–0·731) | 0·715 (0·708–0·721) | 0·698 (0·693–0·703) | 0·677 (0·673–0·680) |
| Multinomial logistic regression (splines): O:E / slope / ICI (%) | 1·11 / 0·98 / 0·15 | 1·04 / 1·03 / 0·42 | 1·02 / 1·03 / 0·65 | 1·00 / 1·05 / 1·16 |
| Random forest: O:E / slope / ICI (%) | 0·75 / 1·11 / 0·52 | 0·91 / 1·12 / 0·92 | 0·96 / 1·10 / 0·96 | 0·98 / 1·15 / 1·62 |
| Histogram gradient boosting: O:E / slope / ICI (%) | 1·09 / 1·03 / 0·12 | 1·03 / 1·02 / 0·22 | 1·01 / 1·00 / 0·23 | 0·99 / 1·01 / 0·27 |
| XGBoost: O:E / slope / ICI (%) | 1·09 / 1·03 / 0·12 | 1·03 / 1·02 / 0·24 | 1·01 / 1·01 / 0·24 | 0·99 / 1·02 / 0·37 |
| LightGBM: O:E / slope / ICI (%) | 1·08 / 1·05 / 0·12 | 1·02 / 1·03 / 0·26 | 1·01 / 1·01 / 0·24 | 0·99 / 1·02 / 0·35 |
| ΔAUC, RF vs regression | −0·004 (−0·007 to −0·001) | −0·002 (−0·004 to −0·001) | −0·003 (−0·004 to −0·001) | −0·002 (−0·003 to −0·001) |
| ΔAUC, HGB vs regression | −0·002 (−0·004 to 0·001) | 0·000 (−0·001 to 0·001) | 0·000 (0·000–0·001) | 0·000 (0·000–0·001) |
| ΔAUC, XGBoost vs regression | 0·000 (−0·002 to 0·002) | 0·001 (0·000–0·002) | 0·001 (0·001–0·002) | 0·001 (0·001–0·002) |
| ΔAUC, LGBM vs regression | −0·001 (−0·003 to 0·001) | 0·001 (0·000–0·002) | 0·001 (0·000–0·002) | 0·001 (0·001–0·002) |

**eTable 2b. Time-dependent AUC (95% CI) and calibration: prostate cancer-specific mortality**

| | 1 year | 3 years | 5 years | 10 years |
|---|---|---|---|---|
| Multinomial logistic regression (splines) | 0·931 (0·920–0·940) | 0·933 (0·927–0·938) | 0·916 (0·911–0·920) | 0·869 (0·864–0·873) |

| | 1 year | 3 years | 5 years | 10 years |
|---|---|---|---|---|
| Random forest | 0·935 (0·925–0·944) | 0·936 (0·931–0·941) | 0·918 (0·914–0·923) | 0·870 (0·866–0·875) |
| Histogram gradient boosting | 0·935 (0·924–0·944) | 0·936 (0·930–0·940) | 0·919 (0·914–0·923) | 0·871 (0·866–0·876) |
| XGBoost | 0·936 (0·926–0·945) | 0·936 (0·931–0·941) | 0·919 (0·915–0·924) | 0·872 (0·867–0·877) |
| LightGBM | 0·936 (0·925–0·945) | 0·936 (0·931–0·941) | 0·919 (0·915–0·924) | 0·872 (0·867–0·876) |
| UCSF-CAPRA | 0·888 (0·876–0·899) | 0·892 (0·885–0·897) | 0·878 (0·873–0·883) | 0·838 (0·834–0·844) |
| NCCN risk group | 0·833 (0·823–0·843) | 0·838 (0·833–0·843) | 0·828 (0·823–0·833) | 0·798 (0·793–0·803) |
| Cambridge Prognostic Group | 0·874 (0·862–0·886) | 0·882 (0·876–0·888) | 0·869 (0·863–0·874) | 0·831 (0·826–0·836) |
| D'Amico risk group | 0·820 (0·810–0·830) | 0·827 (0·821–0·832) | 0·815 (0·811–0·821) | 0·787 (0·783–0·792) |
| ISUP grade group | 0·849 (0·835–0·862) | 0·861 (0·854–0·868) | 0·849 (0·843–0·855) | 0·812 (0·807–0·818) |
| Multinomial logistic regression (splines): O:E / slope / ICI (%) | 1·12 / 0·94 / 0·08 | 1·05 / 1·02 / 0·22 | 1·03 / 1·04 / 0·36 | 1·02 / 1·07 / 0·58 |
| Random forest: O:E / slope / ICI (%) | 0·78 / 1·21 / 0·22 | 0·95 / 1·19 / 0·53 | 0·98 / 1·14 / 0·58 | 1·00 / 1·12 / 0·71 |
| Histogram gradient boosting: O:E / slope / ICI (%) | 1·12 / 1·03 / 0·07 | 1·02 / 1·02 / 0·11 | 1·00 / 1·01 / 0·13 | 1·02 / 1·02 / 0·23 |
| XGBoost: O:E / slope / ICI (%) | 1·11 / 1·04 / 0·07 | 1·02 / 1·04 / 0·13 | 1·00 / 1·02 / 0·11 | 1·01 / 1·03 / 0·24 |
| LightGBM: O:E / slope / ICI (%) | 1·09 / 1·06 / 0·07 | 1·01 / 1·04 / 0·13 | 0·99 / 1·02 / 0·12 | 1·01 / 1·03 / 0·29 |
| ΔAUC, RF vs regression | 0·004 (0·001–0·008) | 0·003 (0·001–0·005) | 0·002 (0·001–0·004) | 0·002 (0·001–0·003) |
| ΔAUC, HGB vs regression | 0·004 (0·001–0·007) | 0·003 (0·002–0·004) | 0·003 (0·001–0·004) | 0·002 (0·001–0·004) |
| ΔAUC, XGBoost vs regression | 0·005 (0·003–0·008) | 0·003 (0·002–0·005) | 0·003 (0·002–0·004) | 0·004 (0·003–0·005) |
| ΔAUC, LGBM vs regression | 0·005 (0·002–0·007) | 0·003 (0·002–0·005) | 0·003 (0·002–0·004) | 0·003 (0·002–0·005) |

**eTable 2c. Time-dependent AUC (95% CI) and calibration: other-cause mortality**

| | 1 year | 3 years | 5 years | 10 years |
|---|---|---|---|---|
| Multinomial logistic regression (splines) | 0·729 (0·715–0·745) | 0·734 (0·727–0·741) | 0·732 (0·726–0·737) | 0·741 (0·738–0·745) |
| Random forest | 0·720 (0·706–0·735) | 0·729 (0·721–0·736) | 0·726 (0·720–0·732) | 0·738 (0·734–0·742) |
| Histogram gradient boosting | 0·726 (0·711–0·742) | 0·733 (0·726–0·741) | 0·731 (0·726–0·737) | 0·741 (0·737–0·745) |
| XGBoost | 0·728 (0·714–0·744) | 0·734 (0·727–0·742) | 0·732 (0·727–0·738) | 0·742 (0·739–0·746) |
| LightGBM | 0·726 (0·711–0·741) | 0·734 (0·727–0·742) | 0·732 (0·727–0·738) | 0·742 (0·738–0·746) |
| UCSF-CAPRA | 0·656 (0·641–0·673) | 0·639 (0·630–0·646) | 0·626 (0·620–0·632) | 0·614 (0·610–0·618) |
| NCCN risk group | 0·634 (0·620–0·648) | 0·616 (0·608–0·623) | 0·601 (0·595–0·607) | 0·593 (0·589–0·597) |
| Cambridge Prognostic Group | 0·643 (0·626–0·659) | 0·627 (0·618–0·635) | 0·613 (0·607–0·619) | 0·602 (0·597–0·605) |
| D'Amico risk group | 0·631 (0·616–0·645) | 0·614 (0·607–0·621) | 0·599 (0·593–0·604) | 0·590 (0·586–0·594) |
| ISUP grade group | 0·618 (0·601–0·636) | 0·604 (0·595–0·613) | 0·594 (0·588–0·600) | 0·585 (0·581–0·589) |
| Multinomial logistic regression (splines): O:E / slope / ICI (%) | 1·10 / 0·98 / 0·08 | 1·03 / 1·01 / 0·16 | 1·02 / 1·01 / 0·26 | 0·99 / 0·99 / 0·74 |
| Random forest: O:E / slope / ICI (%) | 0·72 / 0·97 / 0·32 | 0·88 / 1·05 / 0·44 | 0·95 / 1·08 / 0·51 | 0·98 / 1·15 / 1·23 |
| Histogram gradient boosting: O:E / slope / ICI (%) | 1·07 / 1·01 / 0·06 | 1·03 / 1·01 / 0·13 | 1·02 / 0·99 / 0·15 | 0·98 / 0·98 / 0·33 |
| XGBoost: O:E / slope / ICI (%) | 1·09 / 0·99 / 0·06 | 1·03 / 1·01 / 0·12 | 1·02 / 1·01 / 0·12 | 0·98 / 0·99 / 0·35 |
| LightGBM: O:E / slope / ICI (%) | 1·08 / 1·03 / 0·06 | 1·03 / 1·03 / 0·14 | 1·02 / 1·01 / 0·14 | 0·98 / 1·00 / 0·35 |
| ΔAUC, RF vs regression | −0·009 (−0·013 to −0·004) | −0·005 (−0·008 to −0·003) | −0·006 (−0·007 to −0·004) | −0·004 (−0·005 to −0·003) |
| ΔAUC, HGB vs regression | −0·003 (−0·008 to 0·001) | −0·001 (−0·003 to 0·001) | 0·000 (−0·002 to 0·001) | 0·000 (−0·001 to 0·001) |
| ΔAUC, XGBoost vs regression | −0·001 (−0·004 to 0·002) | 0·001 (−0·001 to 0·002) | 0·001 (0·000–0·002) | 0·001 (0·000–0·002) |
| ΔAUC, LGBM vs regression | −0·003 (−0·006 to 0·000) | 0·000 (−0·001 to 0·002) | 0·001 (−0·001 to 0·002) | 0·001 (0·000–0·001) |

**eTable 3. Temporal validation (development 2010–2012; evaluation 2013–2015): AUC (95% CI) and O:E**

| | 1 year | 3 years | 5 years |
|---|---|---|---|
| ACM: Multinomial logistic regression (splines) | 0·832 (0·818–0·845); O:E 0·95 | 0·823 (0·816–0·831); O:E 0·97 | 0·808 (0·802–0·814); O:E 0·96 |
| ACM: Random forest | 0·824 (0·808–0·838); O:E 0·66 | 0·818 (0·811–0·826); O:E 0·85 | 0·804 (0·798–0·810); O:E 0·89 |
| ACM: Histogram gradient boosting | 0·829 (0·814–0·843); O:E 0·95 | 0·822 (0·815–0·830); O:E 0·96 | 0·807 (0·802–0·813); O:E 0·94 |
| ACM: XGBoost | 0·831 (0·817–0·844); O:E 0·93 | 0·822 (0·816–0·830); O:E 0·96 | 0·807 (0·802–0·813); O:E 0·95 |
| ACM: LightGBM | 0·829 (0·814–0·842); O:E 0·93 | 0·823 (0·816–0·830); O:E 0·95 | 0·807 (0·802–0·813); O:E 0·94 |
| ACM: CAPRA | 0·768 (0·752–0·783) | 0·758 (0·751–0·767) | 0·738 (0·732–0·745) |
| PCSM: Multinomial logistic regression (splines) | 0·945 (0·931–0·956); O:E 0·96 | 0·936 (0·929–0·943); O:E 1·03 | 0·921 (0·915–0·927); O:E 0·98 |

| | 1 year | 3 years | 5 years |
|---|---|---|---|
| PCSM: Random forest | 0·946 (0·933–0·956); O:E 0·72 | 0·937 (0·931–0·944); O:E 0·94 | 0·922 (0·916–0·928); O:E 0·94 |
| PCSM: Histogram gradient boosting | 0·949 (0·937–0·959); O:E 0·97 | 0·939 (0·933–0·946); O:E 1·00 | 0·923 (0·917–0·929); O:E 0·95 |
| PCSM: XGBoost | 0·949 (0·936–0·959); O:E 0·96 | 0·939 (0·932–0·945); O:E 1·01 | 0·923 (0·918–0·929); O:E 0·96 |
| PCSM: LightGBM | 0·949 (0·937–0·959); O:E 0·95 | 0·939 (0·932–0·945); O:E 0·99 | 0·924 (0·918–0·929); O:E 0·95 |
| PCSM: CAPRA | 0·896 (0·882–0·910) | 0·888 (0·879–0·897) | 0·878 (0·870–0·884) |
| OCM: Multinomial logistic regression (splines) | 0·735 (0·713–0·754); O:E 0·95 | 0·729 (0·718–0·742); O:E 0·93 | 0·728 (0·720–0·735); O:E 0·95 |
| OCM: Random forest | 0·720 (0·698–0·741); O:E 0·61 | 0·719 (0·706–0·731); O:E 0·78 | 0·719 (0·711–0·726); O:E 0·87 |
| OCM: Histogram gradient boosting | 0·733 (0·709–0·751); O:E 0·93 | 0·725 (0·713–0·738); O:E 0·93 | 0·726 (0·718–0·733); O:E 0·94 |
| OCM: XGBoost | 0·733 (0·712–0·753); O:E 0·90 | 0·726 (0·715–0·739); O:E 0·92 | 0·726 (0·719–0·734); O:E 0·94 |
| OCM: LightGBM | 0·729 (0·707–0·749); O:E 0·90 | 0·727 (0·715–0·739); O:E 0·92 | 0·726 (0·718–0·734); O:E 0·94 |
| OCM: CAPRA | 0·654 (0·633–0·674) | 0·639 (0·630–0·650) | 0·625 (0·618–0·633) |

**eTable 4. Race-stratified performance of XGBoost: AUC (95% CI); O:E; calibration slope**

| | 1 year | 3 years | 5 years | 10 years |
|---|---|---|---|---|
| ACM: NHW | 0·819 (0·809–0·832); O:E 1·09; slope 1·03 | 0·821 (0·815–0·828); O:E 1·03; slope 1·03 | 0·808 (0·802–0·813); O:E 1·02; slope 1·03 | 0·799 (0·795–0·803); O:E 0·99; slope 1·03 |
| ACM: NHB | 0·775 (0·748–0·801); O:E 1·16; slope 1·00 | 0·783 (0·770–0·795); O:E 1·05; slope 0·99 | 0·777 (0·768–0·786); O:E 1·02; slope 0·98 | 0·762 (0·755–0·769); O:E 0·99; slope 0·94 |
| ACM: Hispanic | 0·851 (0·812–0·889); O:E 0·98; slope 1·08 | 0·843 (0·824–0·859); O:E 1·00; slope 1·05 | 0·816 (0·801–0·829); O:E 0·99; slope 0·99 | 0·802 (0·792–0·811); O:E 0·98; slope 1·01 |
| ACM: NHAPI | 0·793 (0·735–0·842); O:E 1·07; slope 0·99 | 0·795 (0·760–0·824); O:E 0·97; slope 0·97 | 0·792 (0·771–0·811); O:E 0·97; slope 0·96 | 0·795 (0·782–0·807); O:E 0·99; slope 1·04 |
| ACM: NHAIAN | 0·780 (0·666–0·912); O:E 1·53; slope 0·85 | 0·741 (0·636–0·829); O:E 0·92; slope 0·83 | 0·771 (0·706–0·832); O:E 0·94; slope 0·91 | 0·771 (0·723–0·814); O:E 1·05; slope 0·92 |
| PCSM: NHW | 0·945 (0·936–0·955); O:E 1·11; slope 1·06 | 0·941 (0·935–0·946); O:E 1·03; slope 1·05 | 0·923 (0·917–0·928); O:E 1·01; slope 1·03 | 0·873 (0·867–0·878); O:E 1·02; slope 1·03 |
| PCSM: NHB | 0·904 (0·868–0·931); O:E 1·14; slope 0·98 | 0·917 (0·903–0·932); O:E 1·03; slope 1·00 | 0·906 (0·894–0·919); O:E 0·98; slope 1·00 | 0·852 (0·841–0·864); O:E 1·01; slope 0·99 |
| PCSM: Hispanic | 0·937 (0·900–0·965); O:E 1·04; slope 1·06 | 0·937 (0·922–0·951); O:E 1·01; slope 1·03 | 0·920 (0·904–0·933); O:E 1·00; slope 1·00 | 0·887 (0·874–0·896); O:E 1·02; slope 1·04 |
| PCSM: NHAPI | 0·908 (0·852–0·962); O:E 1·12; slope 0·97 | 0·913 (0·886–0·938); O:E 1·00; slope 1·00 | 0·896 (0·871–0·920); O:E 0·95; slope 0·97 | 0·872 (0·855–0·894); O:E 1·01; slope 1·07 |
| PCSM: NHAIAN | 0·971 (0·937–0·996); O:E 1·14; slope 1·27 | 0·960 (0·913–0·990); O:E 0·81; slope 1·25 | 0·952 (0·925–0·978); O:E 0·90; slope 1·19 | 0·882 (0·843–0·921); O:E 1·09; slope 1·00 |
| OCM: NHW | 0·730 (0·710–0·748); O:E 1·08; slope 0·99 | 0·738 (0·730–0·747); O:E 1·03; slope 1·02 | 0·736 (0·730–0·742); O:E 1·02; slope 1·01 | 0·746 (0·741–0·751); O:E 0·98; slope 1·01 |
| OCM: NHB | 0·687 (0·652–0·723); O:E 1·17; slope 0·92 | 0·699 (0·683–0·715); O:E 1·07; slope 0·96 | 0·706 (0·692–0·717); O:E 1·05; slope 0·99 | 0·708 (0·699–0·717); O:E 0·99; slope 0·92 |
| OCM: Hispanic | 0·751 (0·692–0·812); O:E 0·91; slope 1·09 | 0·747 (0·724–0·770); O:E 0·99; slope 1·07 | 0·729 (0·712–0·748); O:E 0·99; slope 0·98 | 0·739 (0·725–0·753); O:E 0·97; slope 0·96 |
| OCM: NHAPI | 0·698 (0·621–0·782); O:E 1·03; slope 0·93 | 0·698 (0·650–0·732); O:E 0·96; slope 0·88 | 0·719 (0·690–0·745); O:E 0·99; slope 0·95 | 0·742 (0·723–0·758); O:E 0·98; slope 1·01 |
| OCM: NHAIAN | 0·708 (0·538–0·889); O:E 1·94; slope 0·90 | 0·598 (0·474–0·732); O:E 1·03; slope 0·49 | 0·621 (0·517–0·715); O:E 0·97; slope 0·61 | 0·679 (0·620–0·732); O:E 1·03; slope 0·79 |

**eTable 5. Race-aware versus race-blind models, 10-year horizon**

| | AUC XGBoost | AUC XGBoost race-blind | O:E XGBoost | O:E XGBoost race-blind | O:E regression | O:E regression race-blind |
|---|---|---|---|---|---|---|
| ACM: NHW | 0·799 (0·795–0·803) | 0·799 (0·795–0·803) | 0·99 (0·98–1·00) | 1·00 (0·99–1·01) | 0·99 (0·98–1·00) | 1·00 (0·99–1·01) |
| ACM: NHB | 0·762 (0·755–0·769) | 0·762 (0·755–0·768) | 0·99 (0·97–1·01) | 1·12 (1·09–1·14) | 1·00 (0·98–1·02) | 1·14 (1·11–1·16) |
| ACM: Hispanic | 0·802 (0·792–0·811) | 0·801 (0·791–0·810) | 0·98 (0·95–1·01) | 0·90 (0·88–0·93) | 1·00 (0·97–1·03) | 0·91 (0·88–0·93) |
| ACM: NHAPI | 0·795 (0·782–0·807) | 0·795 (0·781–0·807) | 0·99 (0·95–1·04) | 0·81 (0·78–0·85) | 1·02 (0·98–1·07) | 0·81 (0·77–0·85) |
| ACM: NHAIAN | 0·771 (0·723–0·814) | 0·771 (0·721–0·812) | 1·05 (0·95–1·15) | 1·08 (0·97–1·18) | 1·00 (0·90–1·10) | 1·07 (0·96–1·17) |

| | AUC XGBoost | AUC XGBoost race-blind | O:E XGBoost | O:E XGBoost race-blind | O:E regression | O:E regression race-blind |
|---|---|---|---|---|---|---|
| PCSM: NHW | 0·873 (0·867–0·878) | 0·873 (0·867–0·878) | 1·02 (1·00–1·04) | 1·03 (1·01–1·05) | 1·02 (1·00–1·04) | 1·04 (1·02–1·07) |
| PCSM: NHB | 0·852 (0·841–0·864) | 0·852 (0·841–0·864) | 1·01 (0·96–1·04) | 1·02 (0·98–1·06) | 1·03 (0·98–1·06) | 1·02 (0·98–1·06) |
| PCSM: Hispanic | 0·887 (0·874–0·896) | 0·887 (0·874–0·896) | 1·02 (0·97–1·06) | 1·01 (0·96–1·06) | 1·05 (0·99–1·10) | 1·02 (0·96–1·07) |
| PCSM: NHAPI | 0·872 (0·855–0·894) | 0·873 (0·855–0·895) | 1·01 (0·93–1·10) | 0·84 (0·78–0·92) | 1·07 (0·99–1·17) | 0·84 (0·78–0·92) |
| PCSM: NHAIAN | 0·882 (0·843–0·921) | 0·884 (0·845–0·923) | 1·09 (0·88–1·29) | 1·12 (0·91–1·33) | 1·03 (0·83–1·22) | 1·21 (0·97–1·45) |
| OCM: NHW | 0·746 (0·741–0·751) | 0·746 (0·741–0·751) | 0·98 (0·97–1·00) | 0·98 (0·97–0·99) | 0·98 (0·97–1·00) | 0·98 (0·97–0·99) |
| OCM: NHB | 0·708 (0·699–0·717) | 0·708 (0·699–0·716) | 0·99 (0·96–1·02) | 1·16 (1·13–1·19) | 0·99 (0·97–1·02) | 1·19 (1·16–1·22) |
| OCM: Hispanic | 0·739 (0·725–0·753) | 0·737 (0·724–0·751) | 0·97 (0·92–1·00) | 0·85 (0·81–0·89) | 0·98 (0·93–1·02) | 0·86 (0·81–0·89) |
| OCM: NHAPI | 0·742 (0·723–0·758) | 0·741 (0·722–0·756) | 0·98 (0·93–1·04) | 0·80 (0·76–0·85) | 1·00 (0·95–1·06) | 0·80 (0·76–0·85) |
| OCM: NHAIAN | 0·679 (0·620–0·732) | 0·680 (0·622–0·735) | 1·03 (0·86–1·22) | 1·05 (0·88–1·24) | 0·99 (0·82–1·15) | 1·00 (0·83–1·17) |

**eTable 6. Treatment-augmented and registry-minimal models: AUC (95% CI)**

| | 1 year | 3 years | 5 years | 10 years |
|---|---|---|---|---|
| ACM: XGBoost | 0·816 (0·806–0·826) | 0·818 (0·813–0·823) | 0·804 (0·800–0·809) | 0·795 (0·792–0·799) |
| ACM: XGBoost + treatment | 0·836 (0·826–0·844) | 0·829 (0·824–0·834) | 0·814 (0·811–0·818) | 0·804 (0·801–0·807) |
| ACM: Regression | 0·816 (0·806–0·827) | 0·817 (0·812–0·822) | 0·803 (0·799–0·808) | 0·794 (0·791–0·798) |
| ACM: Regression + treatment | 0·829 (0·820–0·839) | 0·826 (0·822–0·831) | 0·812 (0·809–0·816) | 0·801 (0·798–0·805) |
| PCSM: XGBoost | 0·936 (0·925–0·945) | 0·936 (0·931–0·942) | 0·919 (0·914–0·924) | 0·872 (0·868–0·877) |
| PCSM: XGBoost + treatment | 0·936 (0·926–0·946) | 0·938 (0·934–0·944) | 0·922 (0·917–0·927) | 0·877 (0·873–0·882) |
| PCSM: Regression | 0·931 (0·920–0·942) | 0·933 (0·927–0·938) | 0·916 (0·910–0·921) | 0·869 (0·864–0·874) |
| PCSM: Regression + treatment | 0·932 (0·920–0·942) | 0·935 (0·930–0·940) | 0·918 (0·913–0·923) | 0·871 (0·867–0·876) |
| OCM: XGBoost | 0·728 (0·713–0·746) | 0·734 (0·727–0·742) | 0·732 (0·727–0·738) | 0·742 (0·739–0·746) |
| OCM: XGBoost + treatment | 0·765 (0·752–0·779) | 0·753 (0·746–0·760) | 0·747 (0·742–0·752) | 0·751 (0·748–0·755) |
| OCM: Regression | 0·729 (0·714–0·745) | 0·734 (0·727–0·742) | 0·732 (0·726–0·737) | 0·741 (0·737–0·745) |
| OCM: Regression + treatment | 0·753 (0·740–0·767) | 0·750 (0·743–0·757) | 0·745 (0·740–0·751) | 0·750 (0·747–0·754) |
| ACM: XGBoost, registry-minimal | 0·806 (0·796–0·818) | 0·807 (0·802–0·813) | 0·794 (0·790–0·798) | 0·789 (0·786–0·793) |
| ACM: Regression, registry-minimal | 0·804 (0·794–0·815) | 0·805 (0·799–0·810) | 0·793 (0·788–0·797) | 0·788 (0·785–0·792) |
| PCSM: XGBoost, registry-minimal | 0·918 (0·907–0·928) | 0·918 (0·913–0·924) | 0·901 (0·895–0·906) | 0·857 (0·853–0·861) |
| PCSM: Regression, registry-minimal | 0·911 (0·898–0·922) | 0·913 (0·907–0·919) | 0·897 (0·891–0·901) | 0·854 (0·849–0·858) |
| OCM: XGBoost, registry-minimal | 0·728 (0·712–0·744) | 0·734 (0·726–0·742) | 0·731 (0·725–0·737) | 0·740 (0·736–0·744) |
| OCM: Regression, registry-minimal | 0·728 (0·712–0·744) | 0·733 (0·726–0·741) | 0·731 (0·725–0·737) | 0·739 (0·735–0·743) |

**eTable 7. Sensitivity analysis: horizon-specific classifiers**

| | AUC | O:E | Slope | Mean predicted | Observed |
|---|---|---|---|---|---|
| 5-year PCSM: Discrete-time competing-risk XGBoost (main) | 0·919 | 1·00 | 1·02 | 3·64% | 3·64% |
| 5-year PCSM: Direct horizon-specific XGBoost (IPCW, competing-risk labels) | 0·918 | 1·00 | 0·99 | 3·64% | 3·64% |
| 5-year PCSM: Naive horizon-specific XGBoost (other-cause deaths excluded) | 0·918 | 0·87 | 0·92 | 4·19% | 3·64% |
| 10-year PCSM: Discrete-time competing-risk XGBoost (main) | 0·872 | 1·01 | 1·03 | 6·85% | 6·94% |
| 10-year PCSM: Direct horizon-specific XGBoost (IPCW, competing-risk labels) | 0·871 | 1·00 | 0·99 | 6·94% | 6·94% |
| 10-year PCSM: Naive horizon-specific XGBoost (other-cause deaths excluded) | 0·869 | 0·73 | 0·83 | 9·45% | 6·94% |

**eTable 8. Cause-specific hazard ratios (95% CI) versus non-Hispanic White men**

| Outcome and group | Model 1 (age) | Model 2 + tumor characteristics | Model 3 (+ social) | Model 4 (+ treatment) |
|---|---|---|---|---|
| PCSM: Non-Hispanic Black | 1·31 (1·24–1·38) | 1·09 (1·03–1·16) | 1·01 (0·95–1·07) | 0·96 (0·91–1·02) |
| PCSM: Hispanic | 1·08 (1·00–1·15) | 0·93 (0·87–0·99) | 0·90 (0·84–0·97) | 0·89 (0·83–0·96) |
| PCSM: Non-Hispanic Asian or Pacific Islander | 0·77 (0·70–0·85) | 0·63 (0·57–0·69) | 0·66 (0·60–0·73) | 0·66 (0·60–0·73) |
| PCSM: Non-Hispanic American Indian or Alaska Native | 1·93 (1·53–2·42) | 1·35 (1·07–1·70) | 1·31 (1·04–1·65) | 1·27 (1·01–1·60) |
| OCM: Non-Hispanic Black | 1·50 (1·45–1·55) | 1·41 (1·37–1·46) | 1·28 (1·24–1·33) | 1·21 (1·17–1·25) |
| OCM: Hispanic | 0·88 (0·84–0·92) | 0·85 (0·81–0·89) | 0·86 (0·82–0·90) | 0·85 (0·81–0·89) |
| OCM: Non-Hispanic Asian or Pacific Islander | 0·70 (0·66–0·74) | 0·67 (0·63–0·71) | 0·75 (0·71–0·80) | 0·76 (0·71–0·81) |
| OCM: Non-Hispanic American Indian or Alaska Native | 1·31 (1·11–1·56) | 1·22 (1·03–1·44) | 1·14 (0·96–1·35) | 1·11 (0·93–1·31) |
| ACM: Non-Hispanic Black | 1·44 (1·40–1·48) | 1·32 (1·28–1·36) | 1·20 (1·16–1·24) | 1·13 (1·10–1·16) |
| ACM: Hispanic | 0·93 (0·90–0·97) | 0·87 (0·84–0·91) | 0·88 (0·84–0·91) | 0·87 (0·83–0·90) |
| ACM: Non-Hispanic Asian or Pacific Islander | 0·72 (0·68–0·76) | 0·65 (0·62–0·69) | 0·72 (0·68–0·76) | 0·73 (0·69–0·76) |
| ACM: Non-Hispanic American Indian or Alaska Native | 1·48 (1·29–1·70) | 1·25 (1·09–1·44) | 1·18 (1·03–1·35) | 1·15 (1·01–1·32) |

Model 4 for other-cause mortality also estimated the following hazard ratios (95% CI): Age (per 10 years), 2·20 (2·15–2·24); Age (quadratic term), 1·03 (1·01–1·04); $Log_2$ PSA, 1·10 (1·09–1·11); Clinical T2, 1·05 (1·02–1·08); Clinical T3–T4, 0·94 (0·87–1·00); cN1, 1·00 (0·92–1·09); M1, 1·00 (0·92–1·09); Positive cores (per 10 percentage points), 1·03 (1·02–1·03); Single, 1·43 (1·37–1·49); Divorced or separated, 1·52 (1·46–1·59); Widowed, 1·38 (1·31–1·46); Unknown marital status, 1·13 (1·09–1·18); Income quartile 1, 1·44 (1·39–1·50); Income quartile 2, 1·15 (1·11–1·20); Income quartile 3, 1·06 (1·01–1·10); Non-metropolitan residence, 1·11 (1·07–1·16); Radical prostatectomy, 0·53 (0·51–0·55); Radiotherapy, 0·87 (0·85–0·90); Chemotherapy, 0·73 (0·60–0·88); Biopsy grade group 2, 1·20 (1·16–1·24); Biopsy grade group 3, 1·24 (1·19–1·29); Biopsy grade group 4, 1·27 (1·22–1·33); Biopsy grade group 5, 1·34 (1·28–1·42).

**eTable 9. Aalen–Johansen cumulative incidence, % (95% CI)**

| | PCSM 5 y | OCM 5 y | PCSM 10 y | OCM 10 y |
|---|---|---|---|---|
| Non-Hispanic White | 3·6% (3·5–3·7) | 6·4% (6·3–6·6) | 6·9% (6·7–7·0) | 17·0% (16·8–17·3) |
| Non-Hispanic Black | 3·9% (3·6–4·2) | 7·7% (7·4–8·1) | 7·5% (7·1–7·8) | 19·6% (19·1–20·1) |
| Hispanic | 3·9% (3·6–4·3) | 5·1% (4·7–5·5) | 7·4% (6·9–7·8) | 14·3% (13·6–14·9) |
| Non-Hispanic Asian or Pacific Islander | 2·9% (2·5–3·3) | 4·7% (4·3–5·2) | 6·2% (5·6–6·8) | 14·0% (13·2–14·9) |
| Non-Hispanic American Indian or Alaska Native | 5·5% (3·4–7·3) | 7·1% (5·0–9·3) | 11·7% (8·6–14·6) | 20·0% (16·4–22·8) |
| NCCN low | 0·4% (0·3–0·4) | 3·9% (3·7–4·1) | 1·4% (1·3–1·5) | 11·8% (11·5–12·1) |
| NCCN intermediate | 0·9% (0·8–0·9) | 6·2% (6·0–6·4) | 2·9% (2·8–3·0) | 17·8% (17·5–18·1) |
| NCCN high | 12·3% (11·9–12·6) | 9·8% (9·5–10·1) | 20·7% (20·2–21·1) | 22·4% (22·0–22·8) |

**eTable 10. Race-aware versus race-blind XGBoost: observed:expected ratio (95% CI) across prespecified strata**

| Group and stratum | Outcome | n | Race-aware | Race-blind |
|---|---|---|---|---|
| Non-Hispanic Black: Age <65 | OCM, 10 y | 12 469 | 1·03 (0·98–1·09) | 1·26 (1·20–1·32) |
| Non-Hispanic Black: Age 65-74 | OCM, 10 y | 6855 | 1·00 (0·96–1·05) | 1·19 (1·13–1·24) |
| Non-Hispanic Black: Age ≥75 | OCM, 10 y | 1644 | 0·95 (0·90–1·00) | 1·02 (0·97–1·08) |
| Non-Hispanic Black: NCCN low | OCM, 10 y | 6296 | 0·97 (0·92–1·04) | 1·21 (1·14–1·29) |
| Non-Hispanic Black: NCCN intermediate | OCM, 10 y | 8782 | 1·03 (0·99–1·07) | 1·21 (1·16–1·26) |
| Non-Hispanic Black: NCCN high | OCM, 10 y | 5890 | 1·00 (0·96–1·05) | 1·13 (1·08–1·18) |
| Non-Hispanic Black: Income Q1 (lowest) | OCM, 10 y | 7824 | 1·01 (0·97–1·05) | 1·16 (1·12–1·22) |
| Non-Hispanic Black: Income Q4 (highest) | OCM, 10 y | 2720 | 1·10 (1·00–1·20) | 1·34 (1·22–1·46) |
| Non-Hispanic Black: Metropolitan | OCM, 10 y | 19 222 | 1·01 (0·98–1·04) | 1·19 (1·15–1·22) |
| Non-Hispanic Black: Non-metropolitan | OCM, 10 y | 1746 | 1·00 (0·93–1·09) | 1·15 (1·07–1·25) |
| Non-Hispanic Black: Married | OCM, 10 y | 10 976 | 1·02 (0·98–1·06) | 1·23 (1·17–1·27) |
| Non-Hispanic Black: Not married | OCM, 10 y | 7604 | 1·00 (0·96–1·04) | 1·15 (1·10–1·19) |
| Non-Hispanic Black: Age <65 | ACM, 10 y | 12 469 | 1·03 (1·00–1·07) | 1·18 (1·14–1·23) |
| Non-Hispanic Black: Age 65-74 | ACM, 10 y | 6855 | 1·01 (0·97–1·04) | 1·15 (1·11–1·19) |
| Non-Hispanic Black: Age ≥75 | ACM, 10 y | 1644 | 0·94 (0·91–0·98) | 1·00 (0·96–1·03) |
| Non-Hispanic Black: NCCN low | ACM, 10 y | 6296 | 0·98 (0·93–1·04) | 1·20 (1·14–1·27) |
| Non-Hispanic Black: NCCN intermediate | ACM, 10 y | 8782 | 1·02 (0·98–1·06) | 1·18 (1·14–1·23) |
| Non-Hispanic Black: NCCN high | ACM, 10 y | 5890 | 1·01 (0·98–1·04) | 1·08 (1·05–1·11) |
| Non-Hispanic Black: Income Q1 (lowest) | ACM, 10 y | 7824 | 1·00 (0·97–1·03) | 1·11 (1·07–1·15) |
| Non-Hispanic Black: Income Q4 (highest) | ACM, 10 y | 2720 | 1·11 (1·03–1·18) | 1·29 (1·19–1·37) |
| Non-Hispanic Black: Metropolitan | ACM, 10 y | 19 222 | 1·01 (0·98–1·03) | 1·14 (1·11–1·16) |

| Group and stratum | Outcome | n | Race-aware | Race-blind |
|---|---|---|---|---|
| Non-Hispanic Black: Non-metropolitan | ACM, 10 y | 1746 | 1·02 (0·96–1·08) | 1·13 (1·06–1·20) |
| Non-Hispanic Black: Married | ACM, 10 y | 10 976 | 1·02 (0·99–1·06) | 1·18 (1·14–1·21) |
| Non-Hispanic Black: Not married | ACM, 10 y | 7604 | 1·01 (0·98–1·03) | 1·11 (1·07–1·14) |
| Non-Hispanic Black: Diagnosed 2010-2012 | OCM, 5 y | 10 167 | 1·10 (1·02–1·16) | 1·29 (1·21–1·37) |
| Non-Hispanic Black: Diagnosed 2013-2015 | OCM, 5 y | 10 801 | 1·00 (0·95–1·07) | 1·18 (1·12–1·27) |
| Non-Hispanic Asian or Pacific Islander: Age <65 | OCM, 10 y | 2767 | 0·94 (0·80–1·10) | 0·74 (0·63–0·86) |
| Non-Hispanic Asian or Pacific Islander: Age 65-74 | OCM, 10 y | 2971 | 0·97 (0·86–1·06) | 0·76 (0·68–0·84) |
| Non-Hispanic Asian or Pacific Islander: Age ≥75 | OCM, 10 y | 1239 | 1·00 (0·92–1·09) | 0·87 (0·79–0·94) |
| Non-Hispanic Asian or Pacific Islander: NCCN low | OCM, 10 y | 2005 | 0·92 (0·79–1·05) | 0·72 (0·62–0·82) |
| Non-Hispanic Asian or Pacific Islander: NCCN intermediate | OCM, 10 y | 2702 | 0·95 (0·86–1·04) | 0·75 (0·68–0·83) |
| Non-Hispanic Asian or Pacific Islander: NCCN high | OCM, 10 y | 2270 | 1·03 (0·94–1·12) | 0·88 (0·81–0·96) |
| Non-Hispanic Asian or Pacific Islander: Income Q1 (lowest) | OCM, 10 y | 365 | 0·70 (0·52–0·93) | 0·54 (0·39–0·71) |
| Non-Hispanic Asian or Pacific Islander: Income Q4 (highest) | OCM, 10 y | 3063 | 1·17 (1·07–1·28) | 0·99 (0·90–1·08) |
| Non-Hispanic Asian or Pacific Islander: Metropolitan | OCM, 10 y | 6823 | 0·98 (0·92–1·04) | 0·80 (0·75–0·85) |
| Non-Hispanic Asian or Pacific Islander: Non-metropolitan | OCM, 10 y | 153 | 0·94 (0·61–1·28) | 0·74 (0·48–1·01) |
| Non-Hispanic Asian or Pacific Islander: Married | OCM, 10 y | 5061 | 0·99 (0·92–1·05) | 0·80 (0·74–0·86) |
| Non-Hispanic Asian or Pacific Islander: Not married | OCM, 10 y | 1011 | 0·94 (0·80–1·08) | 0·77 (0·65–0·89) |
| Non-Hispanic Asian or Pacific Islander: Age <65 | ACM, 10 y | 2767 | 0·98 (0·88–1·08) | 0·79 (0·71–0·87) |
| Non-Hispanic Asian or Pacific Islander: Age 65-74 | ACM, 10 y | 2971 | 0·98 (0·89–1·06) | 0·78 (0·70–0·84) |
| Non-Hispanic Asian or Pacific Islander: Age ≥75 | ACM, 10 y | 1239 | 1·00 (0·94–1·06) | 0·86 (0·80–0·91) |
| Non-Hispanic Asian or Pacific Islander: NCCN low | ACM, 10 y | 2005 | 0·92 (0·79–1·03) | 0·72 (0·62–0·81) |
| Non-Hispanic Asian or Pacific Islander: NCCN intermediate | ACM, 10 y | 2702 | 0·95 (0·85–1·03) | 0·75 (0·68–0·82) |
| Non-Hispanic Asian or Pacific Islander: NCCN high | ACM, 10 y | 2270 | 1·03 (0·98–1·09) | 0·88 (0·83–0·93) |
| Non-Hispanic Asian or Pacific Islander: Income Q1 (lowest) | ACM, 10 y | 365 | 0·84 (0·67–1·03) | 0·67 (0·53–0·81) |
| Non-Hispanic Asian or Pacific Islander: Income Q4 (highest) | ACM, 10 y | 3063 | 1·12 (1·04–1·19) | 0·95 (0·88–1·01) |
| Non-Hispanic Asian or Pacific Islander: Metropolitan | ACM, 10 y | 6823 | 0·99 (0·94–1·04) | 0·81 (0·77–0·85) |
| Non-Hispanic Asian or Pacific Islander: Non-metropolitan | ACM, 10 y | 153 | 0·99 (0·76–1·27) | 0·80 (0·61–1·02) |
| Non-Hispanic Asian or Pacific Islander: Married | ACM, 10 y | 5061 | 0·99 (0·93–1·04) | 0·81 (0·76–0·85) |
| Non-Hispanic Asian or Pacific Islander: Not married | ACM, 10 y | 1011 | 0·98 (0·87–1·07) | 0·81 (0·73–0·89) |
| Non-Hispanic Asian or Pacific Islander: Diagnosed 2010-2012 | OCM, 5 y | 3527 | 1·04 (0·90–1·18) | 0·79 (0·69–0·90) |
| Non-Hispanic Asian or Pacific Islander: Diagnosed 2013-2015 | OCM, 5 y | 3450 | 0·95 (0·81–1·08) | 0·74 (0·63–0·84) |
| Hispanic: Age <65 | OCM, 10 y | 6271 | 0·99 (0·89–1·09) | 0·83 (0·75–0·92) |
| Hispanic: Age 65-74 | OCM, 10 y | 4766 | 0·98 (0·91–1·03) | 0·86 (0·80–0·91) |
| Hispanic: Age ≥75 | OCM, 10 y | 1637 | 0·95 (0·90–1·02) | 0·88 (0·83–0·94) |
| Hispanic: NCCN low | OCM, 10 y | 4397 | 0·94 (0·85–1·02) | 0·83 (0·76–0·91) |
| Hispanic: NCCN intermediate | OCM, 10 y | 4760 | 0·98 (0·91–1·05) | 0·86 (0·81–0·93) |
| Hispanic: NCCN high | OCM, 10 y | 3517 | 0·99 (0·92–1·06) | 0·87 (0·81–0·94) |
| Hispanic: Income Q1 (lowest) | OCM, 10 y | 2312 | 0·95 (0·87–1·04) | 0·80 (0·73–0·88) |
| Hispanic: Income Q4 (highest) | OCM, 10 y | 2744 | 1·02 (0·93–1·13) | 0·98 (0·88–1·08) |
| Hispanic: Metropolitan | OCM, 10 y | 12 183 | 0·98 (0·93–1·02) | 0·87 (0·83–0·91) |
| Hispanic: Non-metropolitan | OCM, 10 y | 487 | 0·86 (0·70–1·03) | 0·73 (0·59–0·88) |
| Hispanic: Married | OCM, 10 y | 8157 | 0·96 (0·90–1·02) | 0·85 (0·80–0·91) |
| Hispanic: Not married | OCM, 10 y | 2813 | 0·97 (0·89–1·06) | 0·83 (0·77–0·91) |
| Hispanic: Age <65 | ACM, 10 y | 6271 | 0·99 (0·92–1·05) | 0·88 (0·82–0·94) |
| Hispanic: Age 65-74 | ACM, 10 y | 4766 | 1·00 (0·95–1·05) | 0·90 (0·86–0·95) |
| Hispanic: Age ≥75 | ACM, 10 y | 1637 | 0·98 (0·94–1·03) | 0·93 (0·89–0·98) |

| Group and stratum | Outcome | n | Race-aware | Race-blind |
|---|---|---|---|---|
| Hispanic: NCCN low | ACM, 10 y | 4397 | 0·93 (0·85–1·02) | 0·83 (0·76–0·91) |
| Hispanic: NCCN intermediate | ACM, 10 y | 4760 | 0·97 (0·91–1·03) | 0·87 (0·82–0·93) |
| Hispanic: NCCN high | ACM, 10 y | 3517 | 1·02 (0·98–1·07) | 0·96 (0·92–1·01) |
| Hispanic: Income Q1 (lowest) | ACM, 10 y | 2312 | 0·97 (0·91–1·03) | 0·86 (0·81–0·91) |
| Hispanic: Income Q4 (highest) | ACM, 10 y | 2744 | 1·04 (0·98–1·12) | 1·02 (0·95–1·09) |
| Hispanic: Metropolitan | ACM, 10 y | 12 183 | 0·99 (0·96–1·02) | 0·91 (0·88–0·94) |
| Hispanic: Non-metropolitan | ACM, 10 y | 487 | 0·92 (0·79–1·06) | 0·82 (0·70–0·94) |
| Hispanic: Married | ACM, 10 y | 8157 | 0·98 (0·94–1·02) | 0·90 (0·86–0·94) |
| Hispanic: Not married | ACM, 10 y | 2813 | 0·99 (0·93–1·05) | 0·90 (0·84–0·95) |
| Hispanic: Diagnosed 2010-2012 | OCM, 5 y | 6425 | 1·04 (0·94–1·14) | 0·88 (0·80–0·96) |
| Hispanic: Diagnosed 2013-2015 | OCM, 5 y | 6249 | 0·94 (0·84–1·04) | 0·80 (0·71–0·88) |

### eMethods 5. Temporal validation in men diagnosed in 2016–2017

The models were refitted on the whole development cohort (2010–2015) and applied unchanged to men diagnosed in 2016–2017, selected with the same criteria. Staging in these years was coded with Derived SEER Combined T, N and M; clinical T and N were accepted only when clinically staged, because pathological T and N come from the surgical specimen, and M was accepted from either source. 32 991 men were included (2253 prostate cancer deaths, 4093 other-cause deaths; median follow-up 6·7 years). Because men with pathological staging were excluded, radical prostatectomy was less common than in the development cohort (1·1% vs 35·8%), so this cohort is not a random sample of later diagnoses. Performance was assessed at 1, 3 and 5 years with the same censoring-weighted measures and 200 bootstrap resamples.

**eTable 11. Temporal validation in men diagnosed in 2016–2017: AUC (95% CI) and 5-year calibration**

| | AUC, 1 year | AUC, 3 years | AUC, 5 years | O:E, 5 years | Slope, 5 years |
|---|---|---|---|---|---|
| PCSM: XGBoost | 0·937 (0·922–0·949) | 0·937 (0·930–0·944) | 0·922 (0·915–0·929) | 0·88 (0·85–0·91) | 0·96 (0·92–0·99) |
| PCSM: Regression | 0·934 (0·919–0·946) | 0·935 (0·927–0·942) | 0·920 (0·912–0·927) | 0·90 (0·87–0·93) | 0·93 (0·91–0·96) |
| PCSM: CAPRA | 0·881 (0·863–0·895) | 0·887 (0·877–0·896) | 0·875 (0·866–0·883) | | |
| PCSM: CPG | 0·858 (0·842–0·871) | 0·872 (0·864–0·879) | 0·860 (0·853–0·866) | | |
| PCSM: NCCN | 0·818 (0·806–0·827) | 0·827 (0·819–0·833) | 0·817 (0·811–0·824) | | |
| OCM: XGBoost | 0·706 (0·680–0·734) | 0·709 (0·696–0·721) | 0·710 (0·699–0·718) | 1·12 (1·08–1·16) | 0·94 (0·89–0·98) |
| OCM: Regression | 0·704 (0·672–0·734) | 0·708 (0·694–0·719) | 0·710 (0·699–0·718) | 1·08 (1·04–1·11) | 0·93 (0·88–0·97) |
| OCM: CAPRA | 0·669 (0·637–0·695) | 0·640 (0·625–0·655) | 0·630 (0·618–0·640) | | |
| OCM: CPG | 0·658 (0·625–0·682) | 0·633 (0·618–0·647) | 0·623 (0·611–0·634) | | |
| OCM: NCCN | 0·641 (0·611–0·666) | 0·626 (0·612–0·639) | 0·617 (0·605–0·628) | | |
| ACM: XGBoost | 0·814 (0·795–0·832) | 0·818 (0·809–0·827) | 0·803 (0·795–0·810) | 1·01 (0·99–1·04) | 0·90 (0·87–0·92) |
| ACM: Regression | 0·813 (0·794–0·830) | 0·817 (0·807–0·825) | 0·802 (0·793–0·809) | 1·00 (0·98–1·02) | 0·88 (0·86–0·91) |
| ACM: CAPRA | 0·770 (0·748–0·789) | 0·763 (0·753–0·772) | 0·745 (0·735–0·753) | | |
| ACM: CPG | 0·754 (0·735–0·771) | 0·751 (0·741–0·760) | 0·734 (0·725–0·742) | | |
| ACM: NCCN | 0·726 (0·707–0·741) | 0·726 (0·716–0·735) | 0·712 (0·703–0·720) | | |

**eTable 12. Race-aware versus race-blind XGBoost in men diagnosed in 2016–2017, 5-year horizon**

| | n | AUC, race-aware | AUC, race-blind | O:E, race-aware | O:E, race-blind |
|---|---|---|---|---|---|
| OCM: Non-Hispanic White | 21 682 | 0·708 (0·696–0·722) | 0·708 (0·696–0·722) | 1·13 (1·09–1·18) | 1·14 (1·10–1·19) |
| OCM: Non-Hispanic Black | 5922 | 0·703 (0·678–0·730) | 0·704 (0·679–0·730) | 1·04 (0·96–1·12) | 1·23 (1·13–1·33) |
| OCM: Hispanic | 3023 | 0·700 (0·665–0·737) | 0·699 (0·663–0·737) | 1·26 (1·10–1·43) | 1·09 (0·95–1·23) |
| OCM: Non-Hispanic Asian or Pacific Islander | 1816 | 0·728 (0·688–0·767) | 0·727 (0·685–0·768) | 1·14 (0·94–1·36) | 0·92 (0·75–1·09) |
| ACM: Non-Hispanic White | 21 682 | 0·801 (0·793–0·810) | 0·802 (0·794–0·811) | 1·01 (0·98–1·04) | 1·03 (1·00–1·06) |

| | n | AUC, race-aware | AUC, race-blind | O:E, race-aware | O:E, race-blind |
|---|---|---|---|---|---|
| ACM: Non-Hispanic Black | 5922 | 0·794 (0·776–0·813) | 0·795 (0·777–0·813) | 0·98 (0·93–1·03) | 1·09 (1·03–1·14) |
| ACM: Hispanic | 3023 | 0·804 (0·779–0·829) | 0·805 (0·779–0·830) | 1·09 (1·00–1·18) | 1·00 (0·92–1·09) |
| ACM: Non-Hispanic Asian or Pacific Islander | 1816 | 0·828 (0·799–0·854) | 0·828 (0·797–0·852) | 1·02 (0·92–1·15) | 0·81 (0·73–0·92) |

Discrimination was maintained. Calibration drifted in the later period (prostate cancer mortality over-predicted, other-cause mortality under-predicted), so absolute O:E values should be interpreted after recalibration; the relative effect of removing race (higher O:E in Black men, lower in Hispanic and Asian or Pacific Islander men) was reproduced.

### eMethods 6. Fixed 10-year binary classification in a held-out test set

Because the 10-year life-expectancy threshold drives the choice of curative treatment, we also framed prediction as binary classification at 10 years: death from any cause within 120 months, and prostate cancer death within 120 months (other deaths coded 0). The cohort was split once at random into training (70%) and test (30%) sets, stratified by outcome status and race. Men with a known 10-year status (died within 120 months or followed for at least 120 months) were analysed (65 899 training, 28 276 test). XGBoost (binary logistic objective, hyperparameters from the primary analysis, early stopping on an inner 10% of the training set) and spline logistic regression were fitted on the training set only. AUROC in the test set used 1000 bootstrap resamples. Risk groups were quartiles of predicted 10-year risk with cut-points from the training set; in all test-set men, cumulative incidence was estimated with the Aalen–Johansen method and hazard ratios versus the lowest quartile with Cox models (cause-specific for prostate cancer death). We used TreeSHAP to describe model predictions in 5000 test-set men. The same pooled out-of-fold predictions from the primary 5-fold analysis gave near-identical AUROCs.

**eTable 13. AUROC (95% CI) in the held-out test set and risk-group hazard ratio**

| | Estimate |
|---|---|
| Death from any cause within 10 years: XGBoost | 0·796 (0·790–0·801) |
| Death from any cause within 10 years: Logistic regression | 0·796 (0·790–0·802) |
| Death from any cause within 10 years: CAPRA | 0·712 (0·705–0·718) |
| Death from any cause within 10 years: CPG | 0·699 (0·692–0·705) |
| Death from any cause within 10 years: NCCN | 0·679 (0·673–0·685) |
| Death from any cause within 10 years: HR, highest vs lowest quartile | 12·08 (11·17–13·06) |
| Prostate cancer death within 10 years: XGBoost | 0·871 (0·863–0·878) |
| Prostate cancer death within 10 years: Logistic regression | 0·870 (0·862–0·877) |
| Prostate cancer death within 10 years: CAPRA | 0·841 (0·832–0·848) |
| Prostate cancer death within 10 years: CPG | 0·835 (0·826–0·843) |
| Prostate cancer death within 10 years: NCCN | 0·799 (0·791–0·806) |
| Prostate cancer death within 10 years: HR, highest vs lowest quartile | 30·64 (25·11–37·37) |

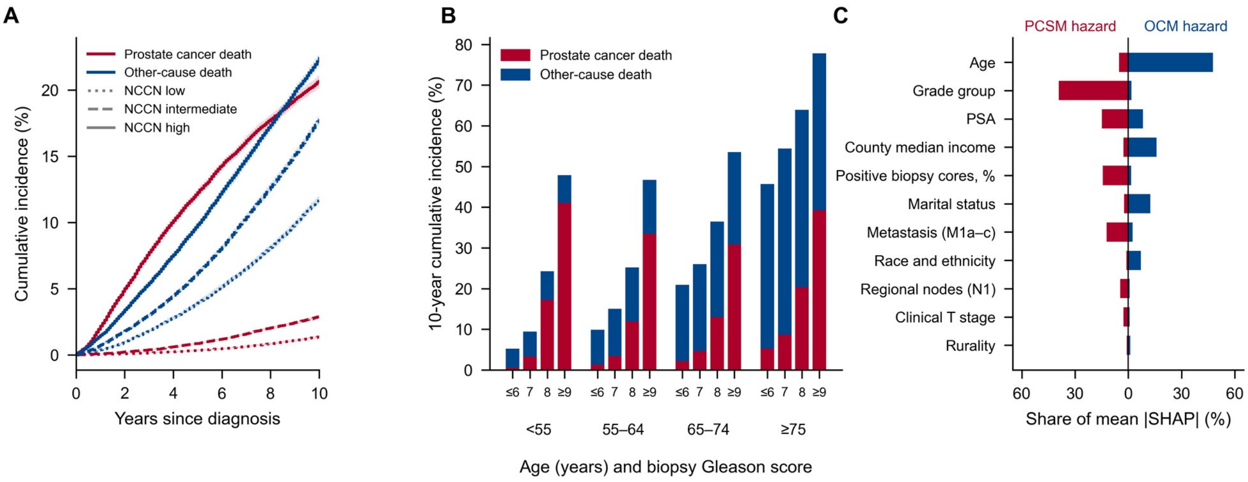


**eFigure 1: Competing-mortality landscape and model explanation** (A) Aalen–Johansen cumulative incidence of prostate cancer (red) and other-cause (blue) death by NCCN risk group, plotted as step functions on the monthly time scale; shaded bands are pointwise bootstrap 95% CIs. (B) 10-year cumulative incidence by age and biopsy Gleason score. (C) Share of mean absolute SHAP value by predictor for the prostate cancer-specific (left) and other-cause (right) hazard in the final XGBoost model (follow-up year 5; 20 000 patients).

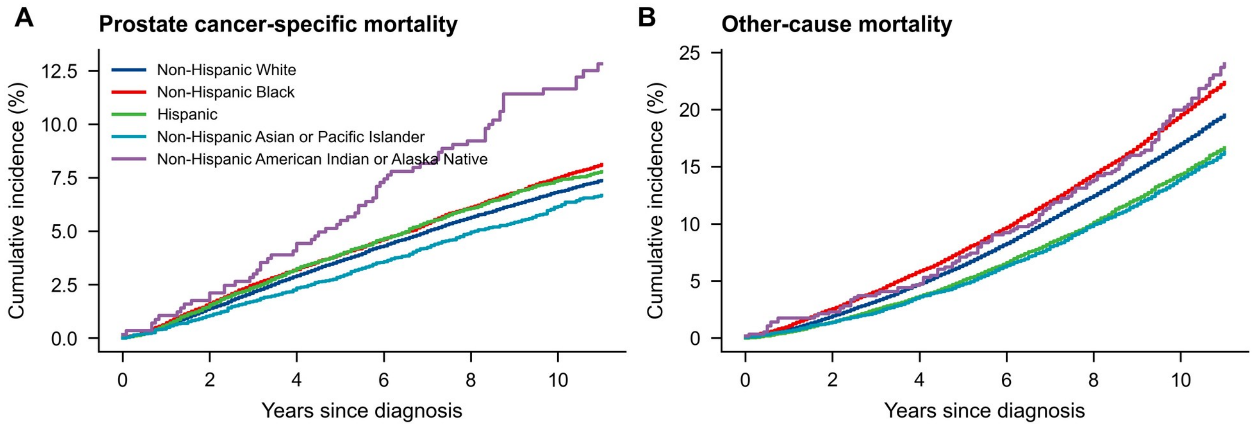


**eFigure 2: Cumulative incidence by race and ethnicity** Aalen–Johansen estimates of (A) prostate cancer-specific and (B) other-cause mortality.

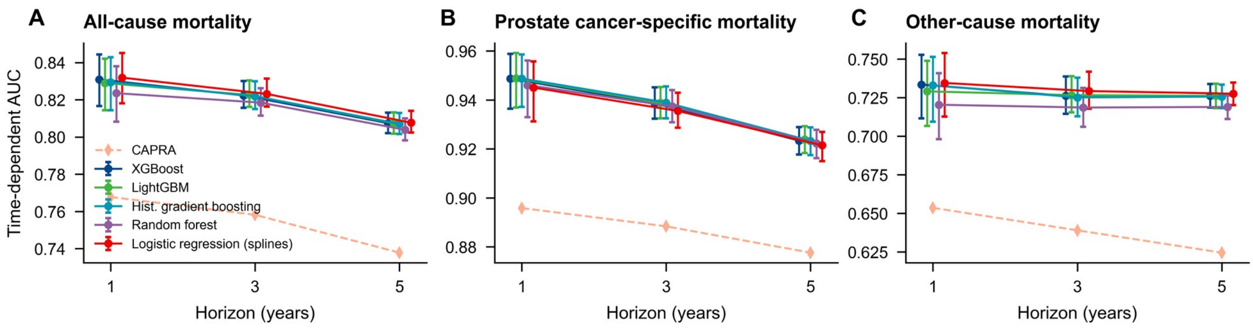


**eFigure 3: Temporal validation** Time-dependent AUC (95% CI) in men diagnosed in the validation period.

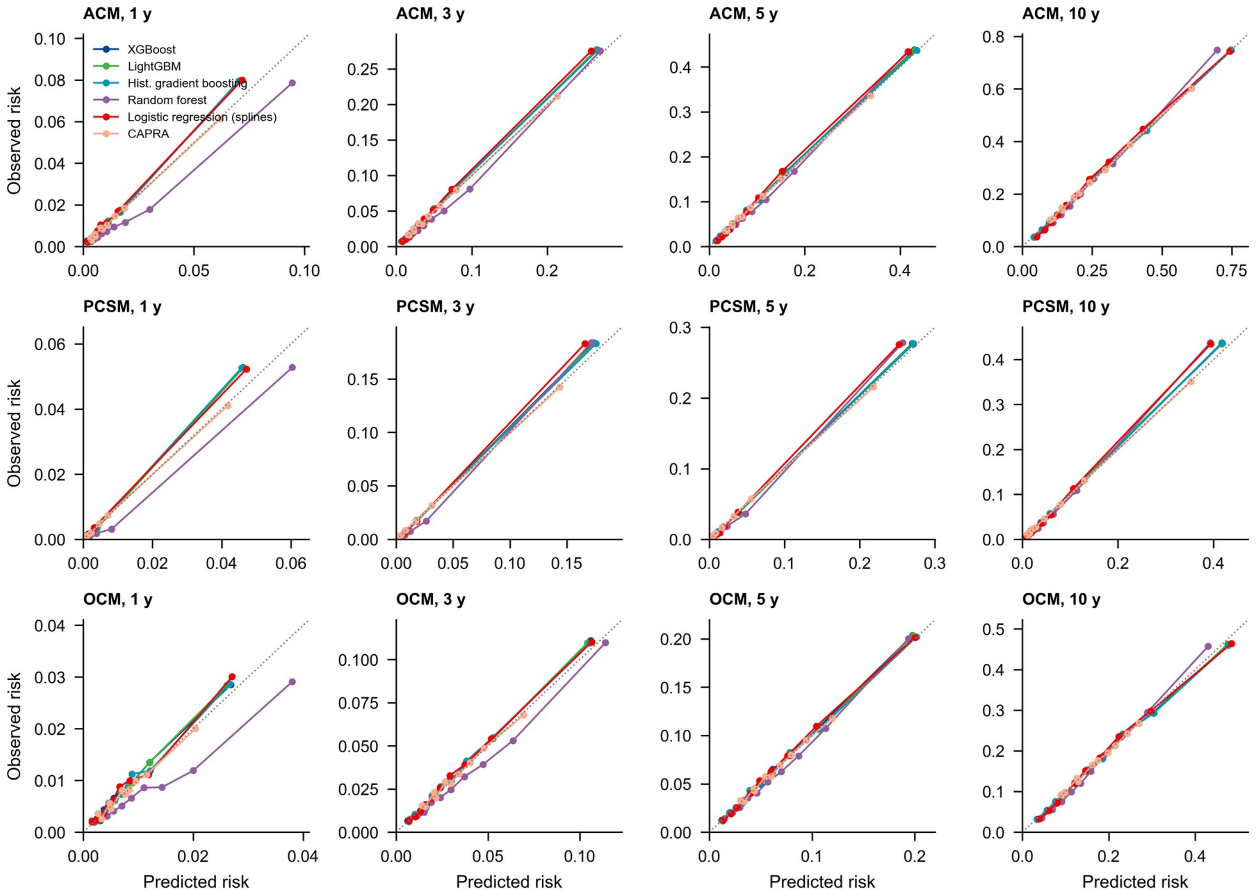


**eFigure 4: Calibration of all models at all horizons** Deciles of predicted versus censoring-weighted observed risk.

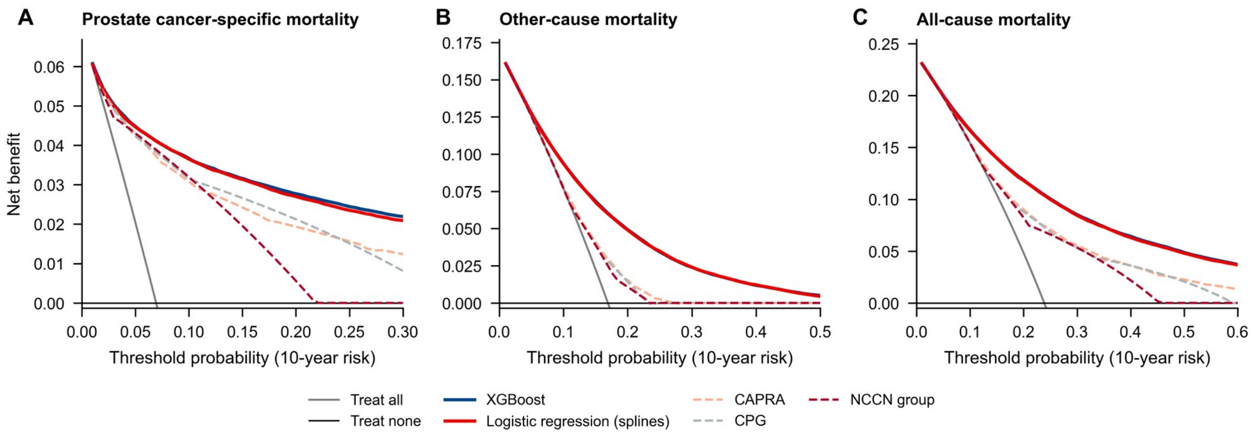


**eFigure 5: Decision-curve analysis for 10-year outcomes** Net benefit of XGBoost, logistic regression and risk-stratification tools (assigned their observed risk per category in training folds) compared with treating all or no men as high risk, for (A) prostate cancer-specific, (B) other-cause and (C) all-cause mortality.

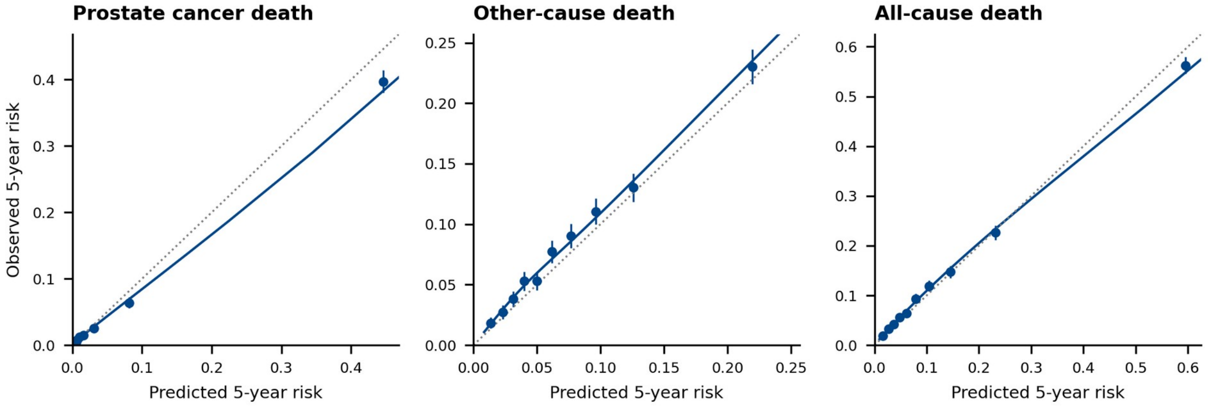


**eFigure 6: Calibration in men diagnosed in 2016–2017** Predicted 5-year risk from XGBoost fitted on 2010–2015 diagnoses versus censoring-weighted observed risk, by decile (points, 95% CI) with a flexible calibration curve (line); dotted line, perfect calibration.

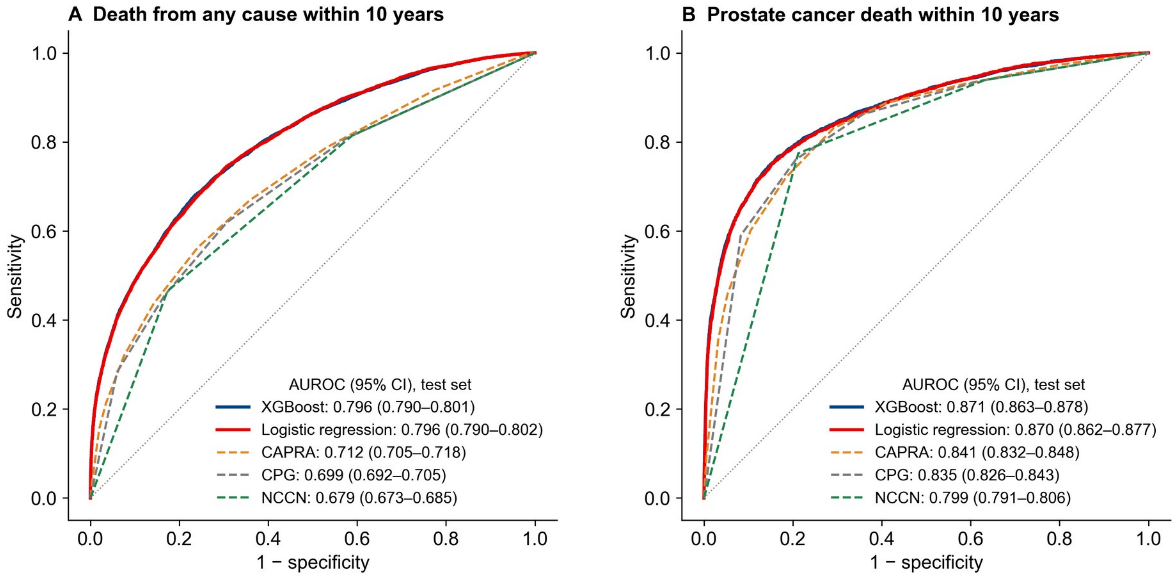


**eFigure 7: ROC curves for 10-year binary outcomes in the held-out test set** AUROC (95% CI) from 1000 bootstrap resamples.

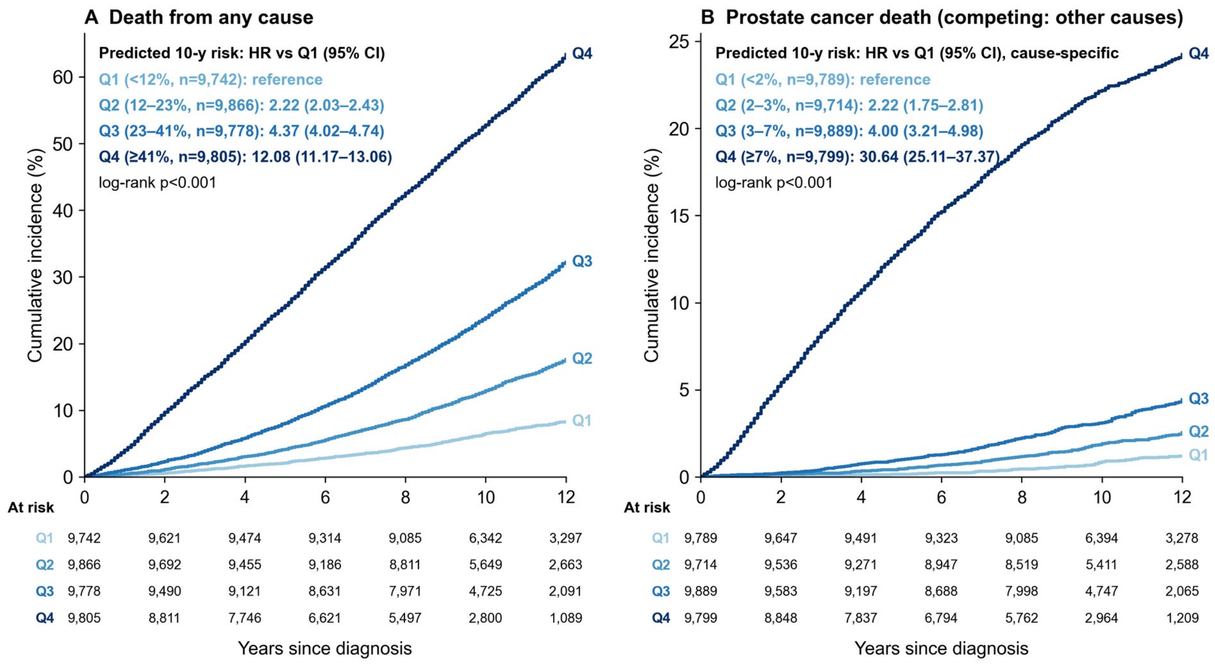


**eFigure 8: Cumulative incidence by quartile of predicted 10-year risk in the test set** Aalen–Johansen estimates; hazard ratios versus Q1 from Cox models (cause-specific for prostate cancer death).

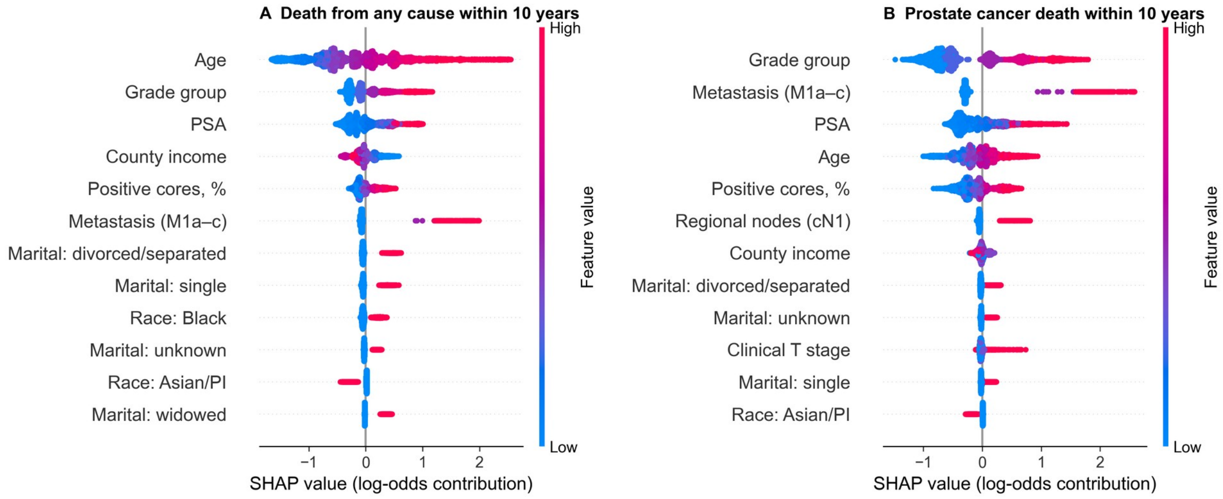


**eFigure 9: SHAP values of the 10-year XGBoost classifiers** Descriptive explanation in 5000 test-set men; positive values increase the predicted risk.

### TRIPOD+AI checklist

**TRIPOD+AI checklist (Collins et al, BMJ 2024) and location in the manuscript**

| Section | Item | Reported in |
|---|---|---|
| Title | 1 | Title page |
| Abstract | 2 | Abstract |
| Background | 3a | Introduction, paragraph 1 |
| | 3b | Introduction, paragraphs 1 and 3 |
| | 3c | Introduction, paragraph 2; Methods, Predictors |
| Objectives | 4 | Introduction, paragraph 3 |
| Data | 5a | Methods, Study design and data source |
| | 5b | Methods, Participants; Study design |
| Participants | 6a | Methods, Study design and data source |
| | 6b | Methods, Participants; figure 1 |
| | 6c | Methods, Predictors (treatment excluded from primary model; secondary model) |
| Data preparation | 7 | Methods, Predictors; appendix eMethods 1–2 |
| Outcome | 8a | Methods, Outcomes |
| | 8b | Methods, Outcomes (registry death certificates; SEER algorithm) |
| | 8c | Not applicable (registry-ascertained outcome) |
| Predictors | 9a | Methods, Predictors |
| | 9b | Methods, Predictors; appendix eMethods 2 |
| | 9c | Not applicable (registry-coded predictors) |
| Sample size | 10 | Methods, Sample size and missing data |
| Missing data | 11 | Methods, Sample size and missing data; eTable 1 |
| Analytical methods | 12a | Methods, Validation and performance measures |
| | 12b | Methods, Model development |
| | 12c | Methods, Model development; eMethods 3 |
| | 12d | Methods, Fairness analyses (subgroup heterogeneity) |
| | 12e | Methods, Validation and performance measures |
| | 12f | Not applicable (no model updating) |
| | 12g | Methods, Model development (CIF computation) |
| Class imbalance | 13 | Methods, Model development (no reweighting or resampling) |
| Fairness | 14 | Methods, Fairness analyses |
| Model output | 15 | Methods, Model development (absolute risks) |
| Training versus evaluation | 16 | Methods, Validation (temporal split) |
| Ethical approval | 17 | Methods, Ethics and consent |
| Funding | 18a | Abstract, Funding |
| Conflicts of interest | 18b | Declaration of interests |
| Protocol | 18c | No protocol link reported |
| Registration | 18d | No registration identifier reported |
| Data sharing | 18e | Data sharing statement |
| Code sharing | 18f | Data sharing statement |
| Patient and public involvement | 19 | Not reported |
| Participants | 20a | Results, paragraph 1; figure 1 |
| | 20b | Table 1 |
| | 20c | eFigure 3; eTable 3 (temporal) |
| Model development | 21 | Results; figure 1; Methods, Sample size |
| Model specification | 22 | Data sharing (code, model files, regression formula) |
| Model performance | 23a | Results; tables 2–3; figures 2–5; eTables 2–5 |
| | 23b | Not applicable (no clustering analysed) |
| Model updating | 24 | Not applicable |
| Interpretation | 25 | Discussion |
| Limitations | 26 | Discussion |
| Usability | 27a | Discussion (inputs are standard clinical staging variables available at diagnosis) |
| | 27b | Discussion |
| | 27c | Discussion (external validation, full-stage data) |
